# Genome-wide DNA methylation, imprinting, and gene expression in human placentas derived from Assisted Reproductive Technology

**DOI:** 10.1101/2023.10.25.23297514

**Authors:** P Auvinen, J Vehviläinen, K Rämö, I Laukkanen, H Marjonen-Lindblad, E Wallén, V Söderström-Anttila, H Kahila, C Hydén-Granskog, T Tuuri, A Tiitinen, N Kaminen-Ahola

**Author notes:** Corresponding author: Nina Kaminen-Ahola. Equal contribution (P Auvinen, J Vehviläinen).

## Abstract

Assisted reproductive technology (ART) has been associated with increased risk for growth disturbance and imprinting disorders, but the molecular mechanisms and whether they are a result of the ART procedures or the underlying subfertility are unknown. Here we performed genome-wide DNA methylation analysis by EPIC Illumina microarrays and gene expression analysis by mRNA sequencing for a total of 80 ART and 77 control placentas, including separate procedure- and sex-specific analyses. ART-associated changes enriched in the pathways of hormonal regulation, insulin resistance, neuronal development, and vascularization. Observed changes in the number of stromal cells as well as *TRIM28* and *NOTCH3* expressions in ART placentas indicated impaired angiogenesis and growth. The enrichment of DNA methylation changes in the imprinted regions and alterations in *TRIM28, ZFP57*, and *NLRP5* suggested defective stabilization of the imprinting. Furthermore, downregulated expression of imprinted endocrine signaling molecule *DLK1*, associated with both ART and subfertility, provides a potential mechanism for the metabolic and phenotypic features associated with ART.

## INTRODUCTION

Approximately one in six couples experience infertility worldwide and the usage of Assisted Reproductive Technology (ART) has increased over the last decades. To date over ten million children have been conceived through these technologies^1^. The main procedure of ART is *in vitro* fertilization (IVF), which may also include intracytoplasmic sperm injection (ICSI), in which a single sperm cell is injected into the oocyte cytoplasm. In addition to the freshly transferred embryos (FRESH), also embryo cryopreservation and frozen embryo transfer (FET) can be a part of IVF or ICSI procedures. Less invasive *in vivo* fertility treatments, such as intrauterine insemination (IUI) and hormone treatments to induce ovulation, are also widely used.

Although children conceived by using the treatments are generally healthy, ART has been associated with increased risks for adverse obstetric and perinatal outcomes such as preterm birth (PTB), low birth weight (LBW), birth defects, and placental anomalies^2,3^. Additionally, long-term effects such as rapid postnatal growth, female early onset puberty as well as cardiovascular and metabolic disorders have been associated with the ART phenotype^4–6^. An increased risk for certain imprinting disorders^7,8^ and a tendency towards higher risk of neurodevelopmental disorders has furthermore been reported^9^. There are method-associated risks, and it has been reported that FRESH derived children have a higher risk for PTB, LBW, and for being small for gestational age (SGA) compared to natural conception. A higher risk of pre-eclampsia and a higher risk for being large for gestational age (LGA) has been noted in pregnancies after FET compared to FRESH^10^. However, it is unclear whether these associations are the result of ART technology *per se* or the underlying subfertility.

The mechanisms by which ART or infertility increases the risk for adverse outcomes are unknown. ART has been suggested to affect fetal development through epigenetic modifications as the procedures take place during extensive epigenetic reprogramming in the gametogenesis and early embryogenesis. Indeed, ART-associated DNA methylation (DNAm) profiles have been reported in several epigenome-wide association studies (EWASs) of human blood^11–18^ and placenta^19–22^. Increasing evidence suggest that the placenta is particularly susceptible to epigenetic changes caused by ART and/or infertility^23–25^ and alterations have been found especially in imprinted genes^19–22^ and also in epigenetically silenced repetitive elements (REs)^26–28^. However, the results of EWASs are not consistent, ART procedure-specific EWASs as well as genome-wide gene expression studies for placentas are scarce, and studies controlled for placental cell type heterogeneity are lacking.

Here, we explored ART-associated genome-wide DNAm by microarrays (Illumina’s Infinium MethylationEPIC) and gene expression by 3’mRNA sequencing (mRNA-seq) of full-term placental samples from ART and naturally conceived singleton newborns (Table 1). The effect of cell type heterogeneity was excluded by adjusting the DNAm analyses by placental cell types. Furthermore, ART-associated phenotypic features were examined. The procedure-specific changes were investigated by analyzing the ART samples in subgroups: newborns derived from IVF or ICSI methods as well as from FRESH or FET procedures. Moreover, to separate the effects of decreased fertility from ART procedures on gene expression, we analyzed placental samples from pregnancies of couples who had went through IUI as well as from subfertile (SF) couples who were about to start or already started ART treatment but got pregnant spontaneously.

**Table 1.**
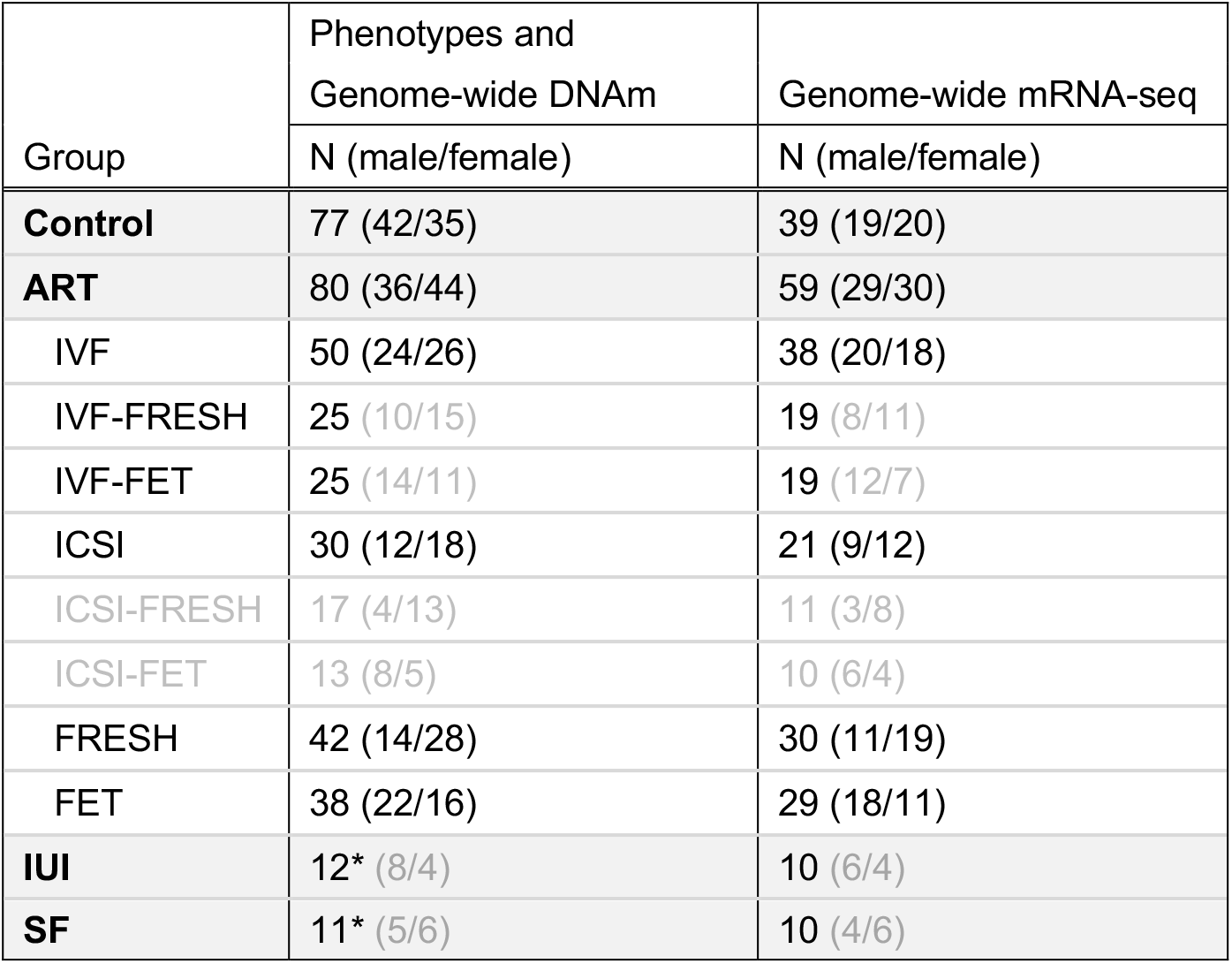
Samples sizes of each study group in phenotype, genome-wide DNAm and mRNA-seq analyses. Study groups are presented in bold with sample sizes in each analysis including the number of samples in the sex-specific analyses. The ART subgroups are shown on a white background. The subgroups or sex-specific samples which are not included in the analyses are written in light gray. IUI and SF groups (indicated with asterisks) are not included in genome-wide DNAm analysis. ART: assisted reproductive technology, IVF: *in vitro* fertilization, ICSI: intracytoplasmic sperm injection, FRESH: fresh embryo transfer, FET: frozen embryo transfer, IUI: insemination, and SF: subfertile.

Males and female fetuses have been shown to differ in terms of placental DNAm^29,30^ as well as in their ability to adapt to adverse intrauterine environments^31,32^. In the ART studies, it has been shown that males have a higher risk to be LGA compared to females^33–35^, but still only a few sex-specific EWASs have been performed so far^12,18,22^. In the current study, we performed sex-specific analyses by examining placentas of male and female newborns separately, including sex chromosomes.

## RESULTS

### Participant characteristics and placental phenotypes

General characteristics of 80 ART and 77 naturally conceived control newborns as well as their mothers were compared. All ART and subgroup-specific information of the phenotype analysis can be found in the Supplementary Table 1. In general, the phenotypes did not differ significantly between ART and control newborns. A total of five SGA (6.3%) (IVF male, two IVF females and two ICSI females, all FRESH) and five LGA (6.3%) (IVF-FRESH, ICSI-FRESH, and ICSI-FET males as well as ICSI-FRESH and ICSI-FET females) newborns were in the ART group, and two SGA newborns (2.6%, male and female) in controls. The gestational age (GA) of ART newborns was significantly shorter, particularly in females (ART: *n* = 44, control: *n* = 35) (*P* = 0.003, *P* = 0.004, respectively), and this was driven by IVF and FRESH newborns (*n* = 50, *n* = 42, respectively). In the ART group, one (1.3%) IVF newborn was preterm (gestational week 36+5). According to the Finnish growth charts^36^, the head circumference (HC) SD (z-score) of ICSI female newborns was significantly smaller compared to control females (*n* = 18, *n* = 35, respectively) (*P* = 0.018). Furthermore, IUI and SF newborns as well as their mothers were compared (*n* = 12, *n* = 11, respectively). The GA of SF newborns was significantly shorter compared to controls (*P* = 0.005) and one female LGA newborn was observed in this group. The mothers of ART newborns were significantly older and they gained less weight during pregnancy compared to the mothers of control newborns (*P* < 0.0001, *P* = 0.002, respectively). Also, the mothers of IUI and SF newborns were significantly older, and the body mass index (BMI) of the IUI mothers was higher compared to controls (*P* = 0.016, *P* = 0.017, *P* = 0.006, respectively).

When the placentas of ART and control newborns were compared, no significant difference in weights was observed (Supplementary Table 1). However, in ART subgroups, FET placentas were significantly heavier compared to FRESH (*n* = 38, *n* = 50, respectively). The DNAm profiles of placental samples and cell type fragment analysis were utilized to determine potential differences between ART and control placentas in five major placental cell types: syncytiotrophoblasts, trophoblasts, stromal cells, Hofbauer cells, and endothelial cells (Supplementary Fig. 1). When comparing the fragments between ART (*n* = 80) and control (*n* = 77) samples, we observed significantly lower number of stromal cells in the ART placentas (*P* = 0.002) (Supplementary Fig. 2) as well as ICSI and FRESH subgroup placentas compared to controls (*n* = 30, *n* = 42, *n* = 77, respectively) (*P* = 0.02 and *P* = 0.03, respectively). Sex-specific analyses revealed significantly lower number of stromal cells in female ART, ICSI, and FRESH placentas compared to control females (*n* = 44, *n* = 18, *n* = 28, *n* = 35, respectively) (*P* = 0.003, *P* = 0.001, *P* = 0.01, respectively) (Supplementary Fig. 2). Furthermore, to exclude the variation caused by different ART procedures, we compared FRESH and FET placentas derived only from the IVF to the controls. Significantly more endothelial cells were observed in the IVF-FRESH placentas compared to the IVF-FET placentas (*n* = 25, *n* = 25, respectively) (*P* = 0.01).

### ART-associated genome-wide DNAm in placenta

To investigate genome-wide average DNAm level (GWAM) in the ART and control placentas (*n* = 80, *n* = 77, respectively), we used the DNAm level of all 746,966 probes in the microarrays (general characteristics in Supplementary Table 1). Furthermore, we examined ART-associated DNAm in REs by comparing the predicted mean DNAm level of CpGs in Alu, endogenous retrovirus (EVR), and long interspersed nuclear element (LINE1) sequences in ART and control placentas. However, we did not observe significant ART-associated alterations in GWAM levels either overall or at any genomic locations relative to gene or CpG island, or in REs (Supplementary Fig. 3 and 4, respectively).

ART-associated genome-wide DNAm was explored by using linear regression model adjusted for cell type, sex, as well as maternal pre-pregnancy BMI and age as covariates. The analysis resulted in 6728 significantly differentially methylated CpG sites (3524 hypo- and 3204 hypermethylated) with FDR < 0.05 (Fig. 1a, Supplementary Table 2). To separate the most prominent changes and to minimize false positive hits, we focused on the CpG sites with DNAm difference of ≥ 5% between ART and control placentas, which are termed as differentially methylated positions (DMPs). There were 164 DMPs associating with 126 genes in ART placentas (FDR < 0.05, Δ*β* ≤ −0.05 and Δ*β* ≥ 0.05), of which 99 were hypo- and 65 hypermethylated. Further, we tested for differentially methylated regions (DMRs) defined as a region with maximal allowed genomic distance of 1000 bp containing three or more CpGs with *P* < 0.05 according to Fisher’s combined probability test. A total of 787 DMRs associating with 687 genes in ART placentas were revealed (Supplementary Table 3).

**Fig. 1.**
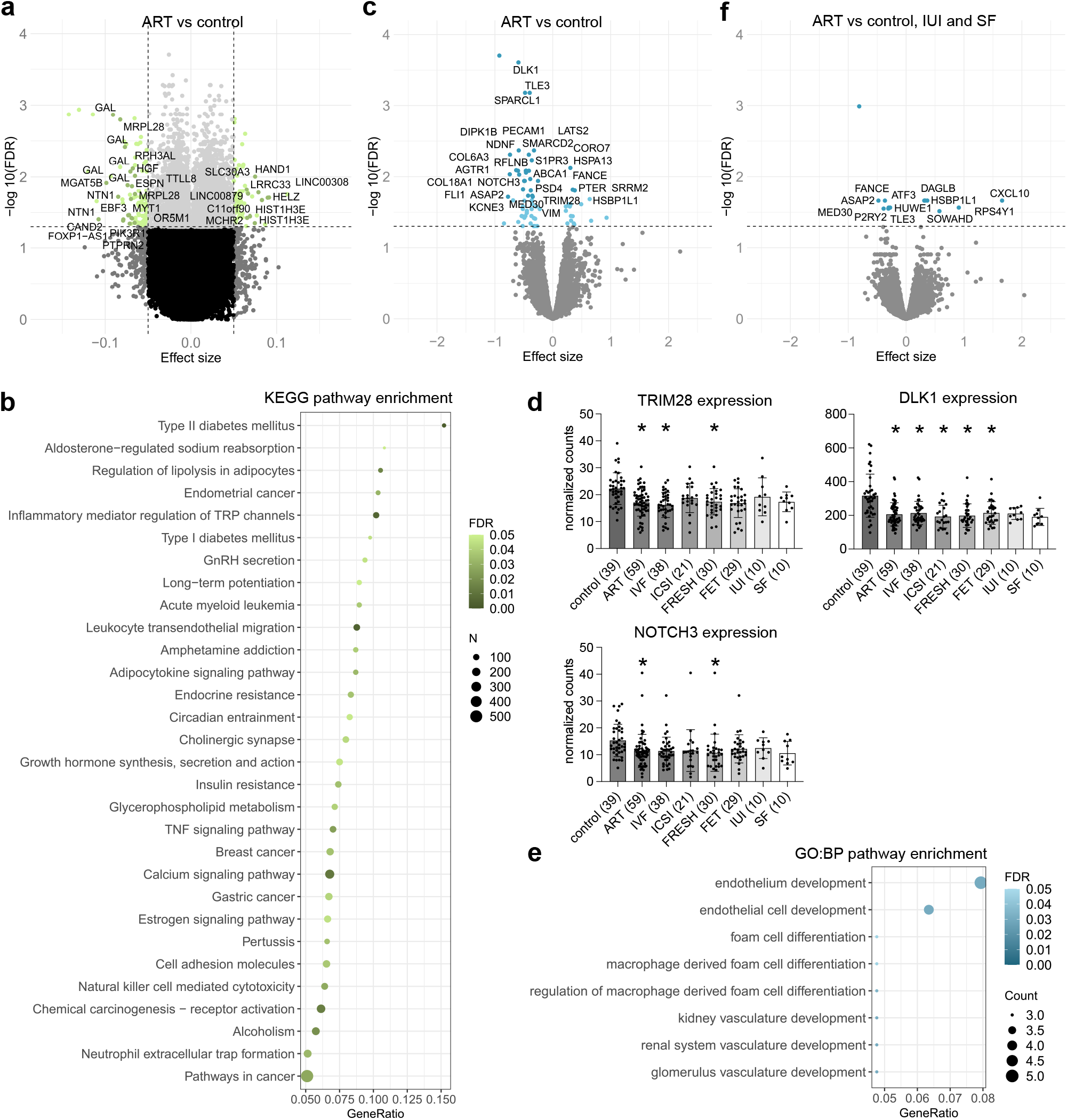
ART-associated differential DNAm and ART-, IUI-, SF-, and *in vitro* culture-associated differential gene expression. **a** Volcano plot showing the distribution of associations between CpG sites and ART. DMPs with Δβ ≤ −0.065 and Δβ ≥ 0.065 are labeled based on UCSC RefGene Name for visualization. **b** Significantly enriched terms identified in KEGG enrichment analysis for DMRs (*P* < 0.05). The 30 most significant pathways are shown. **c** Volcano plot showing the distribution of associations between mRNA expression and ART. The 28 most significant DEGs are labeled for visualization including only confirmed genes. **d** Normalized mRNA-seq counts of *TRIM28, DLK1*, and *NOTCH3* genes in control, ART, IVF, ICSI, FRESH, FET, IUI, and SF placental samples. Data presented as mean ±SD. *FDR < 0.05 in genome-wide mRNA-seq analysis compared to controls. **e** Significantly enriched terms identified in GO:BP enrichment analysis for ART-associated DEGs (FDR-corrected *q*-value < 0.05). All significant pathways are shown. **f** Volcano plot showing the distribution of associations between mRNA expression and *in vitro* culture. All significant DEGs are labeled for visualization including only confirmed genes. In volcano plots, horizontal line marks FDR 0.05 and vertical line marks effect size ±0.05. Control *n* = 77 and ART *n* = 80 in DNAm and control *n* = 39, ART *n* = 59, IVF *n* = 38, ICSI *n* = 21, IUI *n* = 10, and SF *n* = 10 in mRNA-seq analyses.

To get a comprehensive picture on the biological processes (BPs) in which ART-associated DMRs cluster, we performed pathway analyses (Supplementary Table 4). Gene ontology (GO) enrichment analysis of ART-associated DMRs revealed BPs involved in gliogenesis and hormone secretion (*FDR* < 0.05), specifically insulin secretion (*P* < 0.05). When only DMRs located in the regulatory regions (200 and 1500 bp from transcription starting sites (TTSs)) were analyzed, also inflammatory response and cytokine production brought forth (*P* < 0.05). According to KEGG enrichment analysis, ART-associated DMRs linked to terms such as insulin resistance, Type I and II diabetes mellitus (T1D and T2D), growth hormone synthesis, secretion, and action, as well as GnRH secretion, estrogen signaling pathway, and TNF signaling pathway (*P* < 0.05) (Fig. 1b).

Furthermore, placental DMRs in the current study associated with *CHRNE, DUSP22, NECAB3, RASL11B, SNCB,* and *FSCN2*, as well as both DMRs and DMPs with *PCDHGB4, APC2,* and *MSX1,* which all had DNAm changes in the same direction as in the blood samples of ART derived individuals in previous EWASs^14,16,17^. Moreover, *FDFT1, FGF5, MYO7A, TRIM72,* and *C4orf51* have also been observed in a previous study, in which ART placentas were compared to placentas from IUI and SF pregnancies^21^.

### ART-associated genome-wide gene expression in placenta

To study genome-wide ART-associated gene expression, we performed mRNA-seq for ART and control placentas (*n* = 59, *n* = 39, respectively) (general characteristics in Supplementary Table 5). When the mRNA-seq model was adjusted by sex, we observed 71 significantly differentially expressed genes (DEGs) (FDR < 0.05) of which 53 were downregulated and 18 upregulated (Fig. 1c, Supplementary Table 6). One of the most prominently downregulated gene was *delta like non-canonical Notch ligand 1 (DLK1*) (Fig. 1d). This paternally expressed imprinted gene is essential for embryonic development and growth, and its malfunction has been connected to metabolic abnormalities, such as obesity, T2D, and hyperlipidemia in previous human studies^37^. Other interesting downregulated DEGs were *tripartite motif-containing protein 28* (*TRIM28*) and *notch receptor 3* (*NOTCH3)* (Fig. 1d). *TRIM28* is a mediator of epigenetic modifications and its haplosufficiency in mice has been linked to testicular degeneration, age-dependent infertility^38^, and bi-stable epigenetic obesity^39^. Furthermore, *TRIM28* is essential in decidualization and implantation^40^ and is involved in the regulation of female obesity^41^. *NOTCH3,* which is expressed mainly in vascular smooth muscle and pericytes, is needed for the development of fully functional arteries in mouse^42^, and recently it has been reported that *NOTCH3* signaling controls human trophoblast stem cell expansion and proliferation^43^. Interestingly, also ART-associated DMPs and DMRs were observed in its family member *NOTCH1*, which mediates uterine stromal differentiation and is essential for decidualization and implantation in mouse^44^.

When all the placentas were analyzed, moderate correlations were seen between *TRIM28* and *DLK1* as well as between *TRIM28* and *NOTCH3* expressions (*r* = 0.357***, *n* = 118, and *r* = 0.358***, *n* = 118, respectively) (Supplementary Table 7). However, when the control and ART placentas were analyzed separately, correlation between *TRIM28* and *NOTCH3* was only seen in the control (*r* = 0.472*, *n* = 39) but not in the ART placentas. Interestingly, although there is no evidence of direct interaction between *DLK1* and *NOTCH3*, correlations between the expressions of the genes in all, control, ART, and SF placentas were observed (*r* = 0.518***, *n* = 118; *r* = 0.419**, *n* = 39; *r* = 0.427**, *n* = 59; *r* = 0.733*, *n* = 10, respectively).

According to the GO:BP enrichment analysis, ART-associated gene expression is linked predominantly to vasculogenesis including endothelial cell development (*PECAM1, S1PR3, COL18A,* and *ROBO4),* renal vasculature development (*PECAM1, NOTCH3,* and *CD34*), and foam cell differentiation (*AGTR1, ABCA1,* and *CETP*) (FDR-corrected *q*-value < 0.05) (Fig. 1e, Supplementary Table 8). Interestingly, foam cells are a sign for vascular changes in the placenta, the so-called acute atherosis, which has been observed frequently in non-transformed spiral arteries in pregnancies associated with pre-eclampsia, SGA, fetal death, spontaneous PTB, and preterm premature rupture of membranes^45^.

### The separate effects of ART and subfertility

To separate the effects of ART methods and subfertility, we compared gene expression of placentas derived from IUI and SF couples to controls (*n* = 10, *n* = 10, *n* = 39, respectively) (Supplementary Table 9). One DEG, upregulated *TSIX*, a non-coding RNA gene and an antisense to *X Inactive Specific Transcript* (*XIST)*, was observed in IUI and three downregulated DEGs (*ATG9B, SRRM2,* and *INSR*) in SF placentas. When both IUI and SF placentas were compared to controls, five downregulated DEGs (*DLK1, VIM, IGFBP5, TBX2,* and *TRPC6)* were detected. Notably, decreased counts of *DLK1* were also observed in SF placentas (Fig. 1d), suggesting that downregulation of *DLK1* associates with subfertility. By adding IUI and SF samples to the controls we were able to separate the potential effects of hormonal treatments and subfertility from the effects of *in vitro* culture of ART procedures. Interestingly, only 13 DEGs associated with *in vitro* culture (Fig. 1f, Supplementary Table 9).

### ART-associated changes in IVF and ICSI placentas

When IVF and ICSI placentas were compared separately to controls, 140 IVF-associated DMPs linking to 98 genes (Fig. 2a, Supplementary Table 10) and 439 DMRs linking to 417 genes were detected (Supplementary Table 11) (*n* = 50, *n* = 30, *n* = 77, respectively) (general characteristics in Supplementary Table 1). The ICSI placentas associated with 60 DMPs linking to 47 genes (Fig. 2b, Supplementary Table 12) as well as 63 DMRs linking to 48 genes (Supplementary Table 13). Only five DMPs and one gene (*TCN1*) as well as 17 DMR-associated genes were common between the IVF and ICSI placentas and when the IVF placentas were compared to ICSI, none DMPs or DMRs were detected (Fig. 2c, Supplementary Table 14). Although the IVF-associated DMRs and the pathways in which they clustered were similar to ART result (Supplementary Table 15), this separate comparison revealed ICSI-associated DMRs which linked to genes *COLGALT1, DGKZ, PAX8, PAX8-AS1,* and *PDE4B* (Fig. 2c, Supplementary Table 14).

**Fig. 2.**
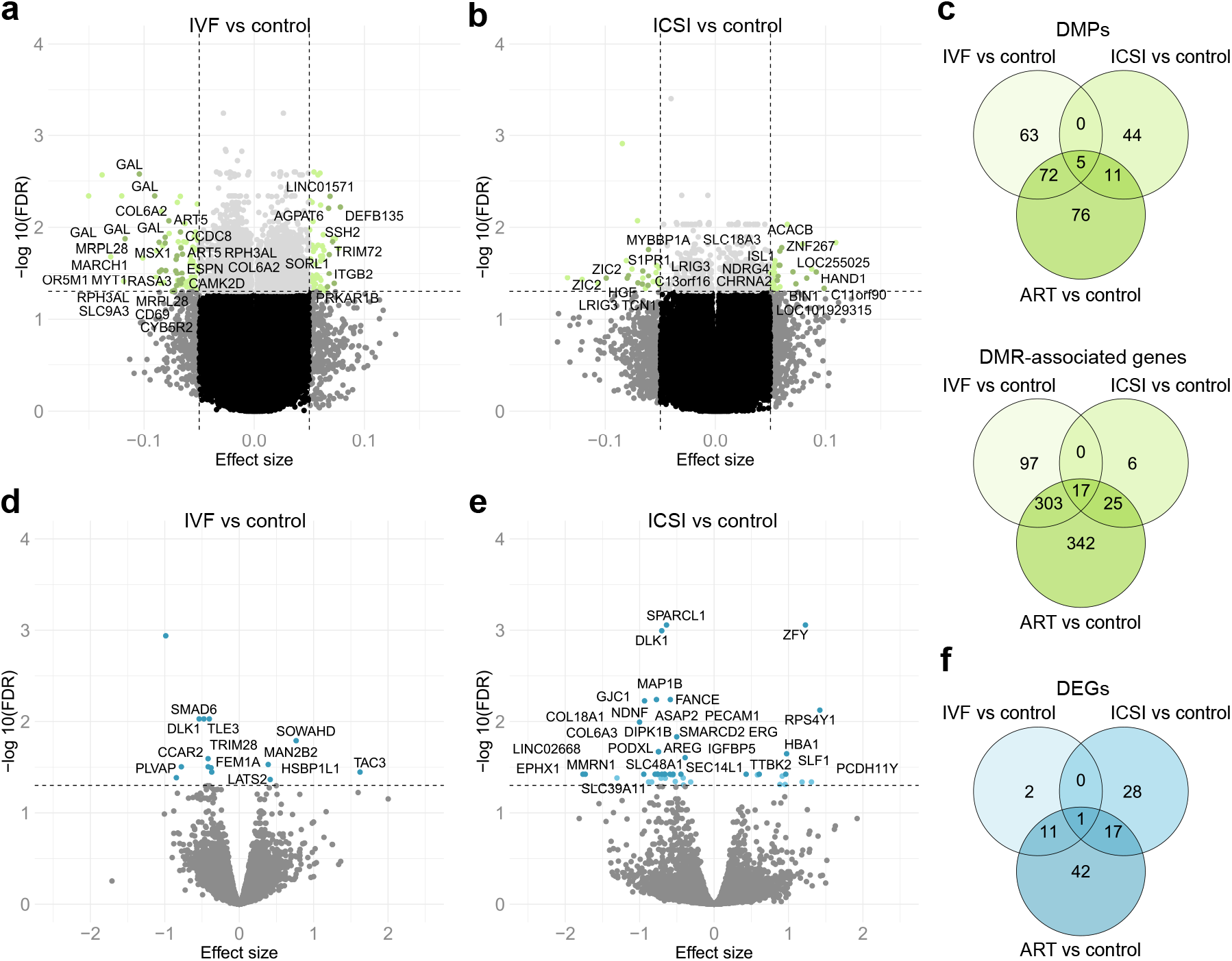
IVF- and ICSI-associated differential DNAm and gene expression. **a** Volcano plot showing the distribution of associations between CpG sites and IVF. DMPs with Δβ ≤ −0.065 and Δβ ≥ 0.065 are labeled based on UCSC RefGene Name for visualization. **b** Volcano plot showing the distribution of associations between CpG sites and ICSI. DMPs with Δβ ≤ −0.055 and Δβ ≥ 0.055 are labeled based on UCSC RefGene Name for visualization. **c** Venn diagram showing the number of ART-associated DMPs as well as DMR-associated genes, which are in common between IVF and ICSI placentas. **d** Volcano plot showing the distribution of associations between mRNA expression and IVF. All significant DEGs are labeled for visualization including only confirmed genes. **e** Volcano plot showing the distribution of associations between mRNA expression and ICSI. The 28 most significant DEGs are labeled for visualization. **f** Venn diagram showing the number of ART-associated DEGs, which are in common between IVF and ICSI placentas. In volcano plots, horizontal line marks FDR 0.05 and vertical line marks effect size ±0.05. Control *n* = 77, IVF *n* = 50, and ICSI *n* = 30 in DNAm and control *n* = 39, IVF *n* = 38, and ICSI *n* = 21 in mRNA-seq analyses.

mRNA-seq analysis for the IVF (*n* = 38) and control (*n* = 39) placentas showed 14 DEGs (10 down- and 4 upregulated), whereas a higher number, 46 genes (34 down- and 12 upregulated), was observed in the ICSI placentas (*n* = 21) (Fig. 2d,f, Supplementary Tables 14 and 16). *DLK1* was the only common DEG between IVF and ICSI placentas. Interestingly, the upregulated genes *ZFY, RPS4Y1, PCDH11Y,* and *DDX3Y* in the ICSI placentas locate on chromosome Y and the expressions of them all correlated significantly with each other in ART male placentas, but not in the controls (Supplementary Table 7). Since male infertility was a common reason (44.4%) for the infertility among the analyzed ICSI male samples (Supplementary Table 17) the upregulated genes *DDX3Y*^46^, *ZFY*^47^, and *PCDH11Y*^48^, which all have been associated with male infertility in previous studies, are notable. Furthermore, the ICSI-associated DEGs *MAP1B, HBA1, EPHX1, HBB*, and *HBG2* enriched in response-to-toxic-substance pathway are also interesting (FDR-corrected *q*-value < 0.05), considering that environmental exposures have been associated with male infertility in previous studies (Supplementary Table 18). Epoxide hydrolases, such as *EPHX1*, are involved in detoxifying and excreting the environmental chemicals which are associated with decreased semen quality and male infertility^49^.

### ART-associated changes in FRESH and FET placentas

Nearly equal amount of DMPs associated with the FRESH and FET placentas when they were compared separately to the controls (*n* = 42, *n* = 38, *n* = 77, respectively): 99 FRESH-associated DMPs were annotated to 97 genes and 93 FET DMPs to 59 genes (Fig. 3a-c, Supplementary Tables 19−21). Furthermore, 253 FRESH- and 100 FET-associated DMRs were observed (FDR < 0.05) (Fig. 3c, Supplementary Tables 21−23). GO enrichment analysis of FRESH DMRs revealed BPs involved in pathways such as cell adhesion via plasma membrane adhesion molecules (FDR < 0.05) and regulation of cytoskeleton organization (*P* < 0.05), whereas FET DMRs clustered to the regulation of astrocyte differentiation and ion transmembrane transporter activity, as well as peptidyl-arginine ADP-ribosylation (*P* < 0.05) (Supplementary Tables 24 and 25, respectively). Interestingly, FRESH DMRs in KEGG enrichment analysis clustered in T2D and insulin resistance, which did not emerge in the analysis for FET DMRs.

**Fig. 3.**
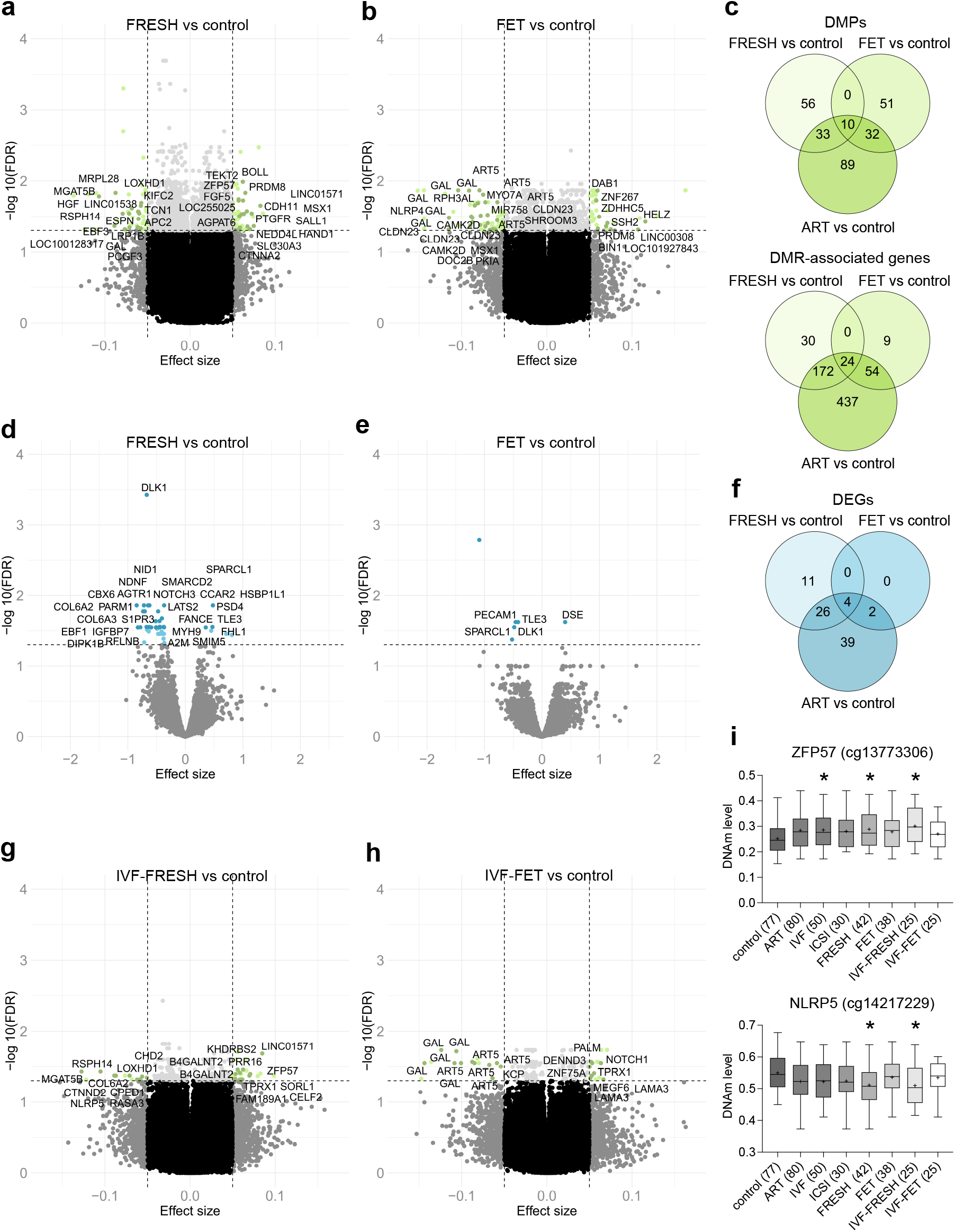
FRESH- and FET-associated differential DNAm and gene expression. **a** Volcano plot showing the distribution of associations between CpG sites and FRESH. DMPs with Δβ ≤ −0.057 and Δβ ≥ 0.057 are labeled based on UCSC RefGene Name for visualization. **b** Volcano plot showing the distribution of associations between CpG sites and FET. DMPs with Δβ ≤ −0.058 and Δβ ≥ 0.058 are labeled based on UCSC RefGene Name for visualization. **c** Venn diagram showing the number of ART-associated DMPs as well as DMR-associated genes, which are in common between FRESH and FET placentas. **d** Volcano plot showing the distribution of associations between mRNA expression and FRESH. The 26 most significant DEGs are labeled for visualization including only confirmed genes. **e** Volcano plot showing the distribution of associations between mRNA expression and FET. All significant DEGs are labeled for visualization including only confirmed genes. **f** Venn diagram showing the number of ART-associated DEGs, which are in common between FRESH and FET placentas. **g** Volcano plot showing the distribution of associations between CpG sites and IVF-FRESH. All DMPs are labeled for visualization. **h** Volcano plot showing the distribution of associations between CpG sites and IVF-FET. All DMPs are labeled for visualization. **i** Normalized DNAm levels of DMPs in *ZFP57* (cg13773306) and *NLRP5* (cg14217229) in placentas of control, ART, and ART subgroups. *FDR < 0.05 in genome-wide DNAm analyses compared to controls. In volcano plots, horizontal line marks FDR 0.05 and vertical line marks effect size ±0.05. Control *n* = 77, ART *n* = 80, IVF *n* = 50, ICSI *n* = 30, FRESH *n* = 42, FET *n* = 38, IVF-FRESH *n* = 25, and IVF-FET *n* = 25 in DNAm and control *n* = 39, FRESH *n* = 30, and FET *n* = 29 in mRNA-seq analyses.

mRNA-seq analysis for the FRESH, FET, and control (*n* = 30, *n* = 29, *n* = 39, respectively) placentas showed 41 FRESH-associated DEGs (35 down- and six upregulated) and only six FET DEGs (five down- and one upregulated) (Fig 3d-f, Supplementary Tables 21 and 26). Downregulated *DLK1* came up from both FRESH and FET placentas, but *NOTCH3* and *TRIM28* were significantly downregulated only in FRESH placentas (Fig. 3d-f). The only upregulated FET-associated DEG was *dermatan sulfate epimerase* (*DSE*), which overexpression has been observed in several cancers and which associates with active angiogenesis, invasion, and proliferation^50^.

Finally, we compared FRESH and FET placentas derived only from the IVF procedure to the controls (*n* = 25, *n* = 25, *n* = 77, respectively). A total of 34 DMPs and 25 DMRs were associating with IVF-FRESH placentas, whereas 29 DMPs and 30 DMRs associated with IVF-FET (Fig. 3g,h, Supplementary Tables 27 and 28, respectively). In both FRESH and IVF-FRESH placentas, the most interesting DMPs located in the regulatory regions of *ZFP57 zinc finger protein* (*ZFP57*) (cg13773306) and *NLR family pyrin domain containing 5* (*NLRP5*) (cg14217229), which have been associated with multilocus imprinting disturbance^51^ (Fig. 3i). Interestingly, *ZFP57,* which also had a DMR in ART, IVF, FRESH, and IVF-FRESH placentas, is a cofactor of *TRIM28*, and they both are required for the germline imprinting^52,53^. The DNAm level of *ZFP57* DMP (cg13773306) correlated with *TRIM28* expression in ART, ICSI and FET placentas, (*r* = 0.33*, *n* = 59; *r* = 0.6**, *n* = 21; *r* = 0.41*, *n* = 38, respectively), with *DLK1* expression in FET placentas (*r* = 0.4*, n = 38), and with *NOTCH3* expression in ICSI placentas (*r* = 0.53*, *n* = 30), supporting associations between DNAm and expression of these genes (Supplementary Table 29). Furthermore, the DNAm level of *NRLP5* DMP (cg14217229) correlated with *NOTCH3* expression in ART, ICSI, and FRESH placentas (*r* = 0.34**, *n* = 59; *r* = 0.54*, *n* = 21; *r* = 0.44*, *n* = 30, respectively). There were only one DEG in IVF-FRESH placentas, downregulated *CBX6* (log fold change = −0.97, FDR = 0.01), and no DEGs in the IVF-FET placentas were observed (*n* = 19, *n* = 19, *n* = 39, respectively).

### Sex-specific differences in ART-derived placentas

To identify sex-specific effects of ART, we performed DNAm analyses including sex chromosomes for the placentas of male and female newborns separately. The GWAM overall level was higher in male placentas compared to females, which is consistent with a previous study^29^ (Supplementary Fig. 3). A higher GWAM overall level was observed in ICSI male placentas compared to control males, and significantly higher compared to IVF males (*n* = 12, *n* = 42, *n* = 24, respectively) (*P* = 0.04). Furthermore, ICSI male placentas had significantly higher GWAM level at genomic locations 5’UTR and S_Shore compared to control males as well as at 5’UTR, S_Shelf and S_Shore compared to IVF males (*P* = 0.03, *P* = 0.03, *P* < 0.05, *P* = 0.01, *P* = 0.03, respectively). By contrast to the males, differences in GWAM overall level were not observed between control, IVF, and ICSI female placentas (*n* = 35, *n* = 26, *n* = 18, respectively). However, the ICSI female placentas had significantly lower GWAM at S_Shelf compared to IVF females (*P* = 0.007).

Moreover, a trend of higher GWAM in both male and female FET placentas compared to FRESH and controls was observed (Supplementary Fig. 3). In FET male placentas the GWAM level was significantly higher at N_Shelf compared to FRESH males, and in FET female placentas significantly lower at 1stExon compared to FRESH females (*n* = 22, *n* = 14, *n* = 16, *n* = 28, respectively) (*P* = 0.03 and *P* = 0.04, respectively).

Sex-specific examination of Alu, LINE1, and EVR REs revealed significantly lower Alu DNAm in ICSI male placentas (*n* = 12) compared to IVF males (*n* = 24) (Supplementary Fig. 4). Furthermore, ICSI female placentas (*n* = 18) had significantly lower LINE1 DNAm level compared to control females (*n* = 35) (*P* = 0.03 and *P* = 0.04, respectively).

In ART male placentas, there were only one hypermethylated DMP compared to control males (ART: *n* = 36, control: *n* = 42), whereas 20 DMPs and eight DMRs were observed in ART females (ART: *n* = 44, control: *n* = 35) (Supplementary Table 30). Interestingly, mRNA-seq for male placentas (ART: *n* = 29, control: *n* = 19) revealed 21 DEGs (18 down, 3 up) (FDR < 0.05) clustering in pathways of endothelial and epithelial cell development in GO:BP enrichment analysis, and none DEGs in the ART females (ART: *n* = 30, control: *n* = 20) (Supplementary Table 31). In IVF male placentas five DEGs (downregulated *PECAM1, SPARCL1, DLK1,* and *S1PR3* and upregulated *LYPLA1*) were observed compared to control males, and in IVF female placentas 11 DMPs and four DMRs were observed compared to control females (*n* = 20, *n* = 19, *n* = 26, *n* = 35 respectively). Despite of the small sample size in sex-specific analysis of ICSI samples, 16 DEGs were observed in ICSI male placentas (ICSI: *n* = 9, control: *n* = 19), including five upregulated genes, *DDX3Y, EIF1AY, ZFY, RPS4Y1*, and *PCDH11Y*, locating all on chromosome Y. In female placentas (ICSI: *n* = 12, control *n* = 20), six DEGs were observed including downregulated *CASP2* and upregulated *TFPI2*, *CSH1, CGA, PSG2*, and *PSG3*.

When FRESH and FET placentas were analyzed sex-specifically, only one downregulated DEG (*SPARCL),* was observed in FRESH male placentas (FRESH: *n* = 11, control: *n* = 19) and 21 DMPs and 2 DMRs in the FRESH female placentas (FRESH: *n* = 28, control: *n* = 35) (Supplementary Table 30 and 31). In FET male placentas, one DMR (*HTT-AS* 3 CpG) (FET: *n* = 22, control: *n* = 42), and 11 DEGs clustering in GO:BP pathway term protein refolding (*HSPA13, HSPA5*), were observed (FET: *n* = 18, control: *n* = 19).

### Associations between placental gene expression and phenotypes

Potential correlations between *TRIM28*, *NOTCH3*, and *DLK1*, and phenotypes of newborns and their mothers were calculated (Supplementary Table 29). When all samples were examined, there was a weak correlation between placental *NOTCH3* expression and birth weight (BW) (SD) (*r* = 0.272**, *n* = 118), and it was driven by male samples in which the correlations between *NOTCH3* and BW as well as *NOTCH3* and birth length (BL) (SD) were observed (*r* = 0.414**, *n* = 58, and *r* = 0.369**, *n* = 58, respectively). Furthermore, *NOTCH3* expression correlated with the BW and BL (SD) of controls (*r* = 0.41*, *n* = 39, and *r* = 0.386*, *n* = 39, respectively), with BW and BL (SD) and placental weight of ICSI (*r* = 0.441*, *r* = 0.503*, and *r* = 0.463*, *n* = 21, respectively), as well as with BW and BL (SD) of FET newborns (*r* = 0.419**, *n* = 29; *r* = 0.441*, *n* = 29, respectively). The correlation between *NOTCH3* and BL among the ICSI samples was driven by male newborns.

In all male samples there was a negative correlation between *TRIM28* expression and maternal age (*r* = −0.339, *P* = 0.009, *n* = 58) and in all the female samples *TRIM28* correlated weakly with GA (*r* = 0.257*, *n* = 60). Furthermore, *TRIM28* expression correlated negatively with maternal weight gain during pregnancy in the control and ART placentas (*r* = −0.418*, *n* = 37; *r* = −0.295*, *n* = 54, respectively). In the ART placentas it was driven by ICSI, specifically by ICSI female placentas (*r* = −0.596**, *n* = 19; *r* = −0.655*, *n* = 11, respectively). Also, placental *TRIM28* expression correlated negatively with maternal weight gain and positively with maternal BMI in the FRESH placentas (*r* = −0.388*, *n* = 26; *r* = 0.363*, *n* = 30, respectively).

Placental *DLK1* expression correlated with ART female BL (SD) (*r* = 0.394*, *n* = 30) driven by ICSI females (*r* = 0.676*, *n* = 12), and negatively with maternal weight gain during the pregnancy in IVF females (*r* = −0.618*, *n* = 15).

### ART- and subfertility-associated placental imprinting

Finally, we focused on genomic imprinting since imprinting disorders have been associated with ART in several previous studies^54^. Imprinted genes are a group of parent-of-origin monoallelically expressed genes, which are maintained by correctly methylated imprinting control regions (ICRs) established in the germline. A total of 19 ART- and/or ART subgroup-associated DMRs on the microarrays are annotated to the imprinted genes^55^ of which 13 were hypomethylated (ART: *n* = 80, IVF: *n* = 50, ICSI: *n* = 30, FRESH: *n* = 42, FET: *n* = 38, control: *n* = 77) (Supplementary Table 32). Six of these (*BLCAP, KCNQ1, KCNQ1OT1, NNAT, PEG3,* and *ZIM2*) have been associated with ART also in previous EWASs^12,16,21,56^ and were found to be hypomethylated as in the current study. To focus specifically on ICRs, we compared 826 CpGs on ICRs compiled by Ochoa and collagues^57^ between ART and control placentas and observed significant differences in 115 sites (90 hypo- and 25 hypermethylated) (*P* < 0.05) (Supplementary Table 32). After multiple testing correction, 16 CpG sites remained significant (Bonferroni adjusted *P* < 0.05). Furthermore, by using genome-wide DNAm data we examined if the ART-associated DNAm changes were particularly enriched in the ICRs. There were 15 ART-associated CpGs (13 hypo- and 2 hypermethylated) locating in the ICRs (FDR < 0.05) and according to the hypergeometric distribution test, imprinted regions were significantly enriched with DNAm changes (*P* = 0.009).

In the current study, we observed ART-associated hypomethylation in *ZFP57* as well as prominent downregulation of paternally expressed *DLK1,* which locates in a paternally imprinted *DLK1-DIO3* locus. Since *ZFP57* binds to and stabilizes paternally methylated ICR in *DLK1-DIO3* locus in mouse^58^ and *TRIM28* is a cofactor of *ZFP57*^52,53^, we examined ICR DNAm at this locus of ART (IVF: *n* = four, ICSI: *n* = four), SF (*n* = four), and control (*n* = six) placentas by traditional bisulphite sequencing (Fig. 4a). The analyzed ICR sequence is next to the repeated sequence motifs, which contribute to the DNAm of the ICR^58^. Maternal and paternal alleles were distinguished by two single nucleotide polymorphisms (SNPs) (rs1884539 and rs75998174) when possible. Interestingly, although we did not see highly methylated paternal and demethylated maternal allele in this specific ICR sequence, we observed significantly decreased total DNAm in the ICR of SF placentas compared to control (CpG1: *P* = 0.003, CpG2: *P* = 0.007) and ART placentas (CpG1: *P* = 0.002, CpG7: *P* = 0.047) (Fig. 4b, Supplementary Fig. 5). However, we did not observe significantly decreased DNAm in the ART placentas compared to controls, which indicated that the ICR hypomethylation is not the only mechanism which affects the complex *DLK1* regulation^59,60^.

**Fig. 4.**
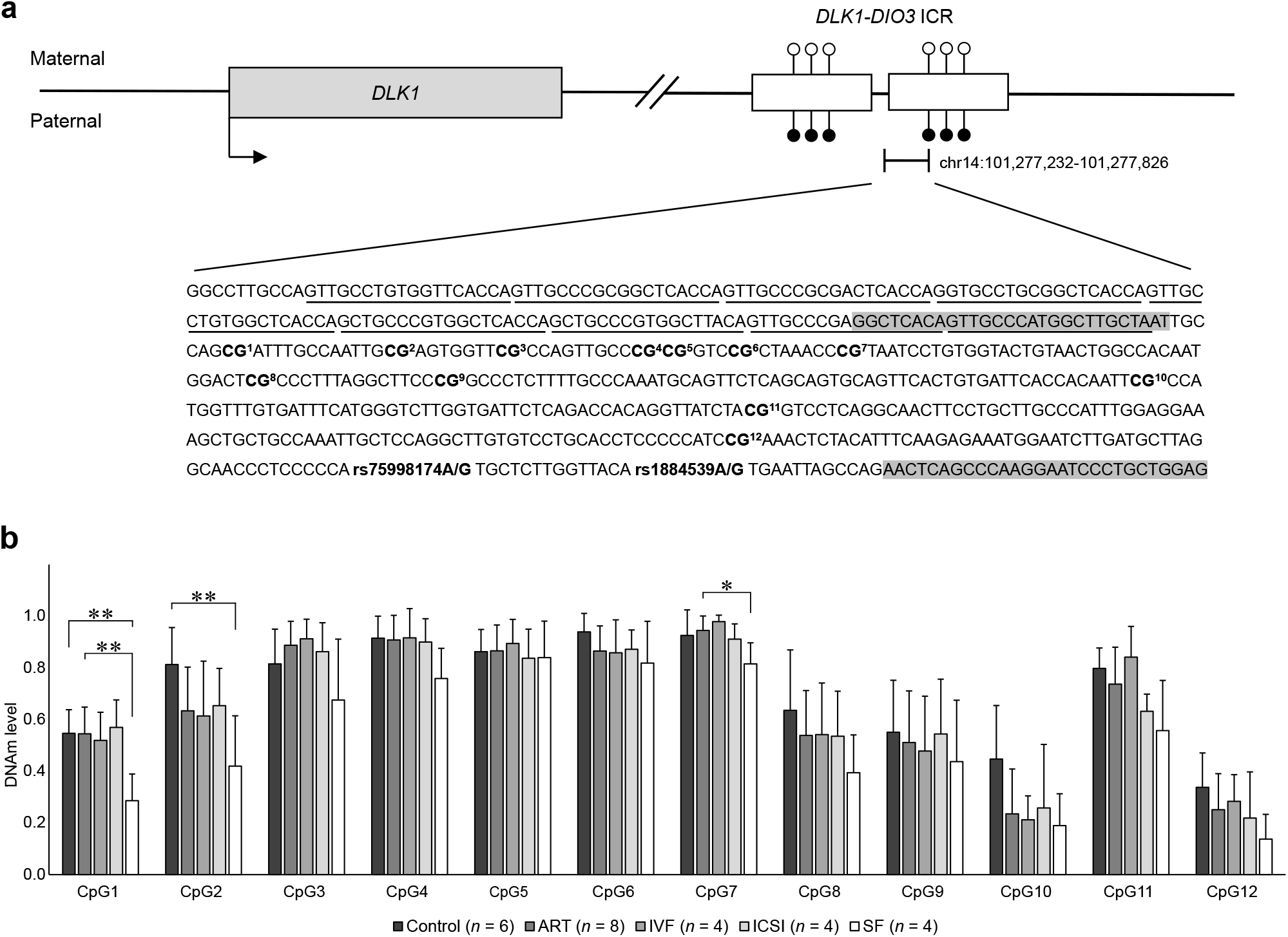
ART- and SF-associated DNAm profiles at *DLK1*-*DIO3* ICR. **a** Schematic figure about *DLK1* and *DLK1-DIO3* ICR. The chromosomal region chr14:101,277,232-101,277,826 at the ICR, containing repeated sequence with nine underlined motifs (chr14:101,277,232-101,277,411) and PCR amplified sequence (chr14:101,277,375-101,277,826), is presented. Primers spanning the amplicon are highlighted in gray and the covered 12 CpG sites and the SNPs (rs1884539A/G and rs75998174A/G) used to distinguish paternal and maternal alleles are bolded. **b** DNAm levels at 12 CpG sites in *DLK1-DIO3* ICR in control (*n* = 6), ART (*n* = 8), IVF (*n* = 4), ICSI (*n* = 4), and SF (*n* = 4) placentas. Differences in DNAm at the CpG sites were calculated between control, ART, and SF placentas. Data presented as mean ±SD. \**P* < 0.05 and \*\**P* < 0.01, One-Way ANOVA followed by Bonferroni post hoc.

## DISCUSSION

In this study, we performed the first genome-wide DNAm and gene expression analyses of human placentas from ART pregnancies as far as we are aware. Those pregnancies are not a homogeneous group, but differ from each other due to the ART procedures, causes of underlying infertility, and sex of the child. By combining the omics data to the phenotypic information and conducting method- and sex-specific analyses, it was possible to perceive the associations between molecular alterations and phenotypic features. There were only a few common ART procedure-associated alterations in the IVF and ICSI placentas, suggesting a strong impact of the underlying sub/infertility on the DNAm and gene expression rather than the effects of the *in vitro* culture. Furthermore, we confirmed previous findings about the procedure-specific effect of FET and observed heavier FET placentas compared to the FRESH placentas. Our observation about the lower amount of gene expression changes in the FET placentas compared to FRESH was consistent with this. Although freezing might be a challenging environmental insult for the embryo, it enables to avoid the hormonal disturbances caused by ovarian stimulation in FRESH. Nearly all (82%) of the FET pregnancies in our study have been started by natural cycle, which could explain the differences between FRESH and FET placentas.

In some studies, the more invasive ICSI procedure has been associated with a higher risk of birth defects^61^, which is concerning due to the increased use of this method. In the current study, more changes in gene expression were observed in the ICSI placentas compared to IVF. Male factor infertility was a prominent reason for ICSI in this study, which could explain the observed alterations. This can be seen in a small number of common ART-associated genes in IVF and ICSI placentas as well as in many male infertility-associated genes on the Y chromosome in the ICSI male placentas. Indeed, in addition to information about the placenta itself, placenta could offer a “molecular window” to early development^62–64^, even to the germ cells. The origin of these early alterations and their potential to provide future biomarkers of infertility should be studied.

Furthermore, in addition to several sex-specific ART-associated alterations, we observed considerable differences between the ICSI males and females. The placental GWAM of ICSI females were more hypomethylated compared to ICSI males, and ICSI female newborns were smaller at birth – even significantly smaller HCs compared to controls were observed. Sex-specific differences in ICSI have been observed also in previous studies. A significantly higher number of trophectoderm cells in ICSI male embryos was detected compared to their female counterparts, which was not observed when IVF male and female embryos were compared^65^. Also previously, ICSI female newborns had significantly smaller BWs (SD) as well as HCs (SD) at the ages of 18 and 36 months (the information about the HC at birth was lacking) compared to controls^66^. The number of sex-specific studies so far is surprisingly small and more studies with larger sample sizes are needed to verify this discovery as well as to find out whether this is caused by the ICSI method or by the underlying male infertility.

In addition to confirming previous ART-associated alterations observed in blood and placental samples, we found new candidate genes *DLK1*, *TRIM28*, and *NOTCH3* for ART-associated phenotypes. By performing cell type fragment analysis, we observed a significantly lower number of stromal cells in the ART placentas compared to controls. Interestingly, it has been suggested that differences in placental mesenchymal stromal cells could lead to impaired vascular development and consequently to restricted growth^67,68^. This is supported by our gene expression analysis, in which the most prominent ART-associated changes were linked to vasculogenesis, and where the downregulation of *NOTCH3* and *TRIM28,* both involved in the regulation of angiogenesis through the VEGFR-DLL4-NOTCH signaling circuit^69^, was observed. Furthermore, *NOTCH3* expression correlated with BW (SD) of control newborns, with BW (SD), BL (SD), and placental weight of ICSI male newborns, as well as BW (SD) and BL (SD) of FET newborns. Since *NOTCH3* and *TRIM28* are not significantly downregulated in placentas of FET newborns − whose size is near to the control newborns − the downregulation of *NOTCH3* could explain the higher risk of LWB and SGA associated with FRESH. Interestingly, among the control and FET groups the maternal hormonal function is not affected: in most cases the embryo was transferred in a natural cycle without the hormonal disturbances caused by ovarian stimulation. The role of hormones brought forth also in the enrichment analysis, where ART-associated DMRs linked to terms such as growth hormone synthesis, secretion, and action, as well as GnRH secretion, and estrogen signaling pathway. Recently, it has been shown that *TRIM28* modulates estrogen receptor α and progesterone receptor signaling, which regulate endometrial cell proliferation, decidualization, implantation, and fetal development^40^. Considering all these observations, the functional links between *NOTCH3, TRIM28*, and the ART-associated cardiovascular phenotype should be studied. Also, the separate effects of hormonal ovarian stimulation in ART protocols and subfertility-associated impaired hormonal function on early development need to be clarified in the future.

Along with *TRIM28* and *NOTCH3*, one of our most interesting discoveries in the ART placentas concerned the prominent downregulation of imprinted *DLK1* as well as the pathways of insulin signaling, insulin resistance, T1D and T2D, which came forth in the DNAm analyses. The developmentally essential *DLK1* is a part of evolutionary conserved Delta-Notch pathway, and it inhibits adipogenesis by controlling the cell fate of adipocyte progenitors^70,71^. According to animal studies, mice without paternally expressed *DLK1* exhibit growth retardation and obesity while *DLK1* overexpression generate decreased fat mass, diet-induced obesity resistance, and reduced insulin signalling^72,73^. In human, mutations in *DLK1* gene have been reported as a cause of central precocious puberty associated with obesity and metabolic syndrome with undetectable DLK1 serum levels^74^. Furthermore, in Temple syndrome, where *DLK1* expression is downregulated due to the maternal uniparental disomy of the imprinted *DLK1* locus, phenotypic features such as prenatal growth failure, short postnatal stature, female early onset puberty, and truncal obesity have been observed^75^. Since increased risks for SGA, rapid postnatal growth, and female early onset puberty^6^ have all been associated with ART children, *DLK1* is a plausible candidate gene for the phenotypic effects associated with ART.

In addition to the downregulation of *DLK1* expression, we observed significant changes in *ZFP57, TRIM28,* and *NLRP5,* which all are needed for the stability of DNAm in the imprinted regions. We observed enrichment of decreased DNAm in several ICRs – common binding sites of *ZFP57* and *TRIM28* – suggesting that the machinery, which normally stabilizes the DNAm of ICRs globally could be impaired. This could explain the high variability in ART-associated imprinted genes observed in previous studies. Previously, mutations or dysregulation of *ZFP57* have been associated with imprinting disorders such as transient neonatal diabetes mellitus^76^, which incidence has been shown to be higher among ART newborns^77^. Furthermore, mutations in *NLRP5* were responsible for early embryonic arrest in infertility patients^78^. More specific examination of *DLK1-DIO3* locus showed decreased DNAm of the ICR in SF placentas. This suggests that the instability of DNAm at this locus associates with subfertility, which is consistent with the decreased *DLK1* expression in the ART, SF, and IUI placentas observed in the current study, and with the previously reported associations between subfertility and decreased placental *DLK1* expression^19^. However, *TRIM28* can regulate the expression of imprinted *DLK1* in several ways^59,60^ and therefore the interaction of these two genes should be examined by functional studies. Also, the effect of ageing on *TRIM28* expression and its associations with both *DLK1* and *NOTCH3* should be studied profoundly, due to the previous association between *TRIM28* and age-dependent infertility in mouse^38^ as well as our observed negative correlation between placental *TRIM28* expression and maternal age.

These findings are also interesting in the perspective of fetal programming. According to the programming hypothesis, adverse conditions such as prenatal malnutrition in the critical developmental period may permanently program physiological processes. If there is plenty of food available postnatally, this programming may prove inappropriate and cause metabolic alterations leading to adult diseases such as obesity, T2D, metabolic syndrome, and cardiovascular disease^79,80^. Previously, Cleaton and colleagues showed in mouse that both *Dlk1* expression in maternal tissues and circulating DLK1 derived from the fetus are necessary for appropriate maternal metabolic adaptations to pregnancy by allowing the switch to fatty acid use when resources are limited during fasting^81^. Also, they reported that the circulating DLK1 level predicts embryonic mass in mouse and lower DLK1 levels were strongly associated with high-resistance patterns of flow in the umbilical artery and slow abdominal circumference growth velocity in a human cohort. Furthermore, DLK1 level enables to differentiate healthy SGA from pathologically small infants. Our observation about the downregulated placental *DLK1* and *TRIM28* as well as their negative correlation with maternal weight gain during the pregnancy suggest that they did not effectively inhibit maternal adipogenesis, resulting in increased maternal weight and impaired maternal metabolic adaptations to the pregnancy. This negative correlation was seen particularly in our IVF and ICSI female as well as FRESH placentas, in the subgroups where the relatively smaller placentas and newborns were derived from. The switch to fatty acid use in maternal tissues during fasting could be impaired, potentially causing restricted fetal growth and a higher risk for adult disorders in the future. However, in the ART-associated phenotype the increased risk for LBW, SGA, rapid postnatal growth, and metabolic disorders would be a consequence of downregulation of *DLK1* caused by subfertility associated disorder, not a consequence of poor maternal nutrition as in the example of fetal programming. Notable, significant associations between subfertility and LBW have been observed in previous studies^82^.

In summary, the observed molecular changes in genome-wide DNAm and gene expression in the ART placentas were relatively subtle, which is a finding in line with the fact that the majority of the ART conceived children are as healthy as newborns from natural conceptions. Decreased expression of *TRIM28*, *DLK1,* and *NOTCH3* as well as ART-associated DNAm in *ZFP57* and *NLRP5* bring forth potential mechanisms for the metabolic and phenotypic features including growth disturbance and imprinting disorders, which have been associated with ART. Considering that the observed changes associated also with subfertility, they offer a precious insight to the molecular background of infertility.

## METHODS

### Epigenetics of ART (epiART) Cohort

Finnish, Caucasian couples applied to fertilization treatment in the Fertility clinic of the Family Federation of Finland or the Reproductive Medicine Unit of Helsinki University Central Hospital, Finland were recruited to this study and the samples were collected during the years 2015–2021. The epiART cohort includes 80 newborns fertilized using ART (IVF *n* = 50 and ICSI *n* = 30) of which 42 using FRESH and 38 using FET embryos. Moreover, the cohort includes newborns from pregnancies of couples who had went through IUI as well as from SF couples who were about to start or already started ART treatment but got pregnant spontaneously. More detailed information about the number of samples and newborns and maternal characteristics is presented in Table 1 and Supplementary Table 1. Parental infertility diagnoses of ART, IUI, and SF samples were categorized into male or female factor infertilities, infertility caused by multiple causes, and unexplained infertility (Supplementary Fig. 6 and Supplementary Table 17). Four cases in the ART group (5%) (IVF 4% and ICSI 3.3%) and one in the IUI group (8.3%) used medication for chronic somatic disease hypothyroidism. Two ART cases (2.5%), both in IVF group (4%), had a psychiatric disorder diagnosis and the other one used medication for it (anxiety disorder). One in the IUI group (8.3%) had medication for a bipolar disorder. Pregnancy related disorders such as gestational diabetes mellitus occurred in seven ART cases (8.8%) (IVF 12%, and ICSI 33.3%), two IUI (16.7%) and two SF (18.2%), hepatogestosis in three ART cases (3.8%) (IVF 4%, ICSI 3.3%) and pregnancy related hypertension in one IUI (8.3%) and one SF (9.1%). A total of 16 ART (20 %) (IVF 22% and ICSI 17%), four IUI (33%), and three SF (27%) deliveries were cesarean sections (CSs). The naturally conceived controls (*n* = 77) were full term neonates born from uncomplicated pregnancies of healthy Finnish, Caucasian mothers collected during the years 2013–2015 in Helsinki University Hospital, Finland. Five (6.5%) of the deliveries were CSs.

Differences in newborn birth measures and maternal characteristics between study groups were calculated, depending on the data distribution, by parametric two-tailed Student’s *t*-test or nonparametric Mann-Whitney U test in the case of two groups, and by parametric One-Way ANOVA followed by Bonferroni post hoc or nonparametric Kruskal-Wallis test followed by pairwise comparisons in the case of multiple groups. Differences in BW, BL, and HC of the newborns were calculated using both anthropometric measures and the SDs of measures based on Finnish growth charts^36^, in which GA at birth, twinning, parity, and gender were considered when calculating the SDs (z-scores) of birth measures. SGA and LGA are characterized by weight and/or length at least 2 SD scores below or above the mean.

### Sample collection and preparation

Placental samples of newborns were collected immediately after delivery. When this was not possible, the placenta was stored in the fridge for a maximum of 12 h and only DNA was extracted for further analyses. The placental biopsies (1 cm^3^) were collected from the fetal side of the placenta within a radius of 2–4 cm from the umbilical cord, rinsed in cold 1× PBS, fixed in RNAlater^®^ (Thermo Fisher Scientific), and stored at −80 °C. Placental genomic DNA was extracted from one to four (3.5 on average) pieces of placental tissue samples using commercial QIAamp Fast DNA Tissue or AllPrep DNA/RNA/miRNA Universal Kits (Qiagen) or standard phenol-chloroform protocol. RNA was extracted from the same placental pieces as DNA (2.8 pieces on average) by AllPrep DNA/RNA/miRNA Universal Kit. RNA quality was assessed using an Agilent 2100 Bioanalyzer (Agilent Technologies, Inc.), which was provided by the Biomedicum Functional Genomics Unit (FuGU) at the Helsinki Institute of Life Science and Biocenter Finland at the University of Helsinki.

### DNAm microarrays

#### DNAm measurements and data processing

Genomic DNA (1000 ng) from available placental samples (sample information in Supplementary Table 1) was sodium bisulfite-converted using the Zymo EZ DNAm™ kit (Zymo Research), and genome-wide DNAm was assessed with Infinium Methylation EPIC BeadChip Kit (Illumina) following a standard protocol at the Institute for Molecular Medicine Finland. The raw DNAm dataset was pre-processed, quality controlled, and filtered using ChAMP R package^83^ with default settings: the detection *P*-value cutoff was set at *P* = 0.01, and probe bead count > 3 in at least 95% of samples. All probes and samples passed these quality control thresholds and were included in the subsequent steps. Type-I and Type-II probes were normalized using the *adjustedDasen* method^84^ in wateRmelon R package^85^. Potential effects caused by technical factors and biological covariates were studied from singular value decomposition plots. The data was corrected for the effects of array, slide, batch, DNA extraction method, and placental type (stored in a fridge before sampling or not) by the Empirical Bayes method using the R package ComBat^86^. After ComBat adjustment, probes located in sex chromosomes and probes binding to polymorphic and off-target sites were filtered. Moreover, probes in Finnish-specific SNPs, which overlap with any known SNPs with global minor allele frequency were removed. Population-specific masking was obtained from Zhou et al.^87^.

Subsequently, a total of 746,966 probes were retained for further downstream analysis. Probes in sex chromosomes (16,945 probes for males and 16,651 probes for females) were included in sex-specific analyses. Annotation information was merged to corresponding probes from IlluminaHumanMethylationEPICanno.ilm10b4.hg19 R package^88^, which is based on the file “MethylationEPIC_v-1-0_B4.csv” from Illumina^89^. Probe genomic locations relative to gene and CpG island were annotated based on University of California, Santa Cruz (UCSC) database. If the location information was missing, probe was marked as “unknown.” In the case of multiple location entries, group “others” was used. Otherwise, the following abbreviations were used: TSS1500: 1500 bp upstream of transcription start site, TSS200: 200 bp upstream of TSS, UTR: untranslated region, N_shelf: north shelf, N_shore: north shore, S_shore: south shore, S_shelf: south shelf. Probes were annotated to genes based on, in addition to UCSC, also GENCODE Basic V12 and Comprehensive V12 databases.

#### DMP analysis

Genome-wide differential DNAm analysis by using M values was performed by R package Limma^90^, and the linear model was adjusted to consider biological covariates newborn sex and maternal age and pre-pregnancy BMI. Planet R package^91^ was used to count placental cell-type fractions by CIBERSORT method with unfiltered data and used as an adjusting factor in the model. β values were used for visualization and interpretation of the results. DMPs were considered as significant when DNAm difference was greater than 5% (Δβ ≤ −0.05 and Δβ ≥ 0.05) and FDR-corrected *P*-value smaller than 0.05. Benjamini-Hochberg procedure was used to control for FDR. To calculate the differences in placental cell type composition between the study groups, depending on the data distribution, parametric or nonparametric two-tailed *t*-test in the case of two groups, and parametric One-Way ANOVA followed by Bonferroni *post hoc* or nonparametric Kruskal-Wallis test followed by Wilcoxon Rank Sum Exact test in the case of multiple groups were used.

#### DMR and Pathway analysis

DMRcate R package^92^ was used to analyze DMRs. The method uses minimum description length for detecting region boundaries in DMR identification. DMRcate was adjusted to determine probes (≥ 3) in a region with maximal allowed genomic distance of 1000 bp having FDR < 0.05. DMRs with Fisher’s combined probability test *P* < 0.05 were considered significant. DMRs were annotated to imprinted genes based on Geneimprint database^55^. Pathway analysis was performed for significant DMRs by *goregion* function in missMethyl R package^93^ and GO:BP and KEGG knowledgebases were used as a source. When the GO terms were not significant after FDR correction, the terms with the nominal *P*-value < 0.05 were reported.

#### GWAM analysis

β values of all probes in the array (normalized, ComBat adjusted, and filtered data) were used to calculate placental GWAM^94^ levels sample-wise. Differences between the study groups were calculated by parametric or nonparametric two-tailed *t*-test depending on the data distribution in the case of two groups. In the case of multiple groups, One-Way ANOVA followed by Tukey’s honestly significant difference (HSD) test was used.

#### RE DNAm analysis

Processed DNAm data (M values) was used to predict DNAm in Alu, EVR, and LINE1 elements sample-wise using Random Forest-based algorithm implemented by REMP R package^95^. Less reliable predicted results were trimmed according to quality score threshold 1.7 and missing rate 0.2 (20%). Differences between the study groups were calculated by parametric or nonparametric two-tailed *t*-test depending on the data distribution in the case of two groups. In the case of multiple groups, One-Way ANOVA followed by Tukey’s HSD test was used.

#### ICR analysis

Enrichment of ART-associated CpGs at ICRs was measured with hypergeometric distribution analysis. In total of 826 CpG sites located at ICRs^57^ were identified in our genome-wide DNAm data, of which 15 CpGs were significantly differentially methylated (FDR < 0.05). Enrichment of these ICR-associated CpGs were compared to all CpGs in the EPIC array with *phyper* function in R. All CpGs at the ICRs were compered between ART and control placentas by two-tailed Student’s *t*-test. Bonferroni adjustment was used for multiple comparison.

### Traditional bisulfite sequencing

A total of eight ART (four IVF, four ICSI, two/sex), four SF (two/sex), and six control placental samples (three/sex) were subjected to bisulfite sequencing. Samples were chosen based on mRNA-seq counts, control samples having the highest and ART and SF samples the lowest counts. Paternal and maternal alleles were distinguished according to rs1884539(A/G) and rs75998174(A/G) polymorphisms at the *DLK1-DIO3* ICR. Primers were designed to incorporate the polymorphisms using Bisulfite Primer Seeker tool (Zymo Research)^96^ (forward 5’GGTTTATAGTTGTTTATGGTTTGTTAAT and reverse 5’CTCCAACAAAAATTCCTTAAACTAAATT). The 451 bp amplicon (chr14:101,277,375-101,277,826) covered 12 CpG sites at the ICR. Two separate bisulfite conversions were performed for 500 ng of genomic DNA using EZ DNA Methylation™ kit (Zymo Research) and pooled afterwards. To avoid possible PCR bias, three independent 20 μl PCR reactions (HotStar PCR kit, Qiagen) were performed per sample using annealing temperature 56 °C. PCR reactions were gel isolated, and the three reactions of each sample were pooled and purified using NucleoSpin Gel and PCR Clean-up Kit (Macherey-Nagel). The purified PCR fragments were ligated into pGEM^®^-T Easy Vector (Promega) and cloned by standard protocol. The recombinant-DNA clones were purified using NucleoSpin^®^ Plasmid EasyPure kit (Macherey-Nagel) according to manufacturer’s instructions. The sequences were analyzed by BIQ Analyzer^97^ excluding the clones with lower than 90% conversion rate from the dataset. A total of eight to 30 clones of each individual were successfully sequenced (Supplementary Fig. 5). DNAm differences in CpG sites between study groups (control, ART, SF) were calculated by One-Way ANOVA followed by Bonferroni post hoc.

### mRNA-seq analysis

#### Differential expression analysis

Drop-seq pipeline^98^ was used to construct the mRNA-seq count table for available placental RNA samples (sample information in Supplementary Table 5) provided by FuGU. A total of 34,989 transcripts were identified for downstream analysis. Principal component analysis implemented in DESeq2^99^ was used to identify batch effects, and ComBat-seq^100^ was used to adjust separate mRNA-seq batches. Differential expression analysis was performed by DESeq2 R package, with model adjusting for newborn sex. Genes were considered as differentially expressed when FDR-corrected *P*-value was < 0.05. Benjamini-Hochberg procedure was used to control for FDR.

#### Pathway analysis

*enrichgo* function in R package clusterProfiler version 4.0^101^ was used to perform gene-set enrichment analysis for significant DEGs. The GO knowledgebase was used as the source for identifying significantly enriched BP terms (FDR-corrected *q*-value < 0.05). Benjamini-Hochberg procedure was used to control for FDR.

### Correlation analysis

The within sample gene expression correlations were calculated using ComBat-seq-adjusted and Transcripts Per Million-normalized mRNA-seq counts and the correlations between gene expression, *ZFP57* (cg13773306), *NLRP5* (cg14217229) DNAm, newborn phenotype, and maternal characteristics using ComBat-seq-adjusted and DESeq2-normalized mRNA-seq counts by Spearman rank correlation.

### Statistical analysis

All statistical analyses were conducted in R versions 4.2.3 and 4.3.1^102^, IBM SPSS Statistics for Windows, version 29.0 (IBM Corp.), or GraphPad Prism 9 software (GraphPad Software, Inc.). All data are expressed as the mean with ±SD for a normal distribution of variables. Statistical analyses were performed as described in the relevant method sections and in the figure legends.

## ABBREVIATIONS

*ART:*: Assisted reproductive technology
*BL:*: Birth length
*BMI: Body mass index BP:*: Biological process
*BW:*: Birth weight
*CS:*: Cesarean section
*DEG:*: Differentially expressed gene
*DLK1:*: Delta like non-canonical Notch ligand 1
*DMP:*: Differentially methylated position
*DMR:*: Differentially methylated region
*DNAm:*: DNA methylation
*DSE:*: Dermatan sulfate epimerase
*EVR:*: Endogenous retrovirus
*EWAS:*: Eepigenome-wide association study
*FET:*: Frozen embryo transfer
*FRESH:*: Fresh embryo transfer
*FuGU:*: Biomedicum Functional Genomics Unit
*GA:*: Gestational age
*GO:*: Gene ontology
*GWAM:*: Genome-wide average DNAm level
*HC:*: Head circumference
*HSD:*: Honestly significant difference
*ICR:*: Imprinting control region
*ICSI:*: Intracytoplasmic sperm injection
*IUI:*: Intrauterine insemination
*IVF:*: In vitro fertilization
*LBW:*: Low birth weight
*LGA:*: Large for gestational age
*LINE1:*: Long interspersed nuclear element
*mRNA-seq:*: 3’mRNA sequencing
*N_shelf:*: north shelf
*N_shore:*: north shore
*NLRP5:*: NLR family pyrin domain containing 5
*NOTHC3:*: Notch receptor 3
*PTB:*: Preterm birth
*RE:*: Repetitive element
*S_shore:*: south shore
*S_shelf:*: south shelf
*SF:*: Subfertile
*SGA:*: Small for gestational age
*SNP:*: Single nucleotide polymorphism
*T1D:*: Type I diabetes mellitus
*T2D:*: Type II diabetes mellitus
*TRIM28:*: Tripartite motif-containing protein 28
*TSS:*: Transcription starting site
*TSS200:*: 200 bp upstream of TSS
*TSS1500:*: 1500 bp upstream of transcription start site
*UCSC:*: University of California, Santa Cruz
*UTR:*: Untranslated region
*XIST:*: X Inactive Specific Transcript
*ZFP57:*: ZFP57 zinc finger protein

## Contributions

P.A., J.V., V.S-A., C.H-G., T.T., A.T. and N.K-A. contributed to the study design. H.K., V.S-A., and A.T. recruited the study participants. P.A., J.V., K.R., E.W., H.M., C.H. and N.K-A. contributed to the sample collection and processing. P.A., K.R., I.L. and N.K-A. contributed to the laboratory experiments. P.A., J.V., K.R., I.L. and N.K-A. contributed to the data analysis. P.A., J.V. and N.K-A. drafted the manuscript. All authors contributed to the revision of the manuscript. All authors gave final approval of the version to be published.

## Supplementary material

Supplementary Figures 1–6

Supplementary Tables 1–32

## Ethics approval and consent to participate

Informed consent was obtained from all participants and the study was approved by the Ethics Committee of Helsinki University Central Hospital (386/13/03/03/2012, 285/13/03/03/2013).

## Availability of data

The datasets supporting the conclusions of the current study are included within the article and its additional files. Due to the sensitive nature of the patient data used, the data sets are not and cannot be made publicly available.

## Acknowledgements

We gratefully thank all families that participated in this study. We would also like to acknowledge Teija Karkkulainen, Riikka Vass, and Ira Larsen for their contribution to the recruitment of the participants as well as Samuli Auvinen for his assistance with bisulphite sequencing visualization.

## REFERENCES

1. European Ivf Monitoring Consortium, f.t.E.S.o.H.R. et al. ART in Europe, 2018: results generated from European registries by ESHRE. Hum Reprod Open 2022, hoac022 (2022).

2. Qin, J.B. et al. Worldwide prevalence of adverse pregnancy outcomes among singleton pregnancies after in vitro fertilization/intracytoplasmic sperm injection: a systematic review and meta-analysis. Arch Gynecol Obstet 295, 285–301 (2017).

3. Vermey, B.G. et al. Are singleton pregnancies after assisted reproduction technology (ART) associated with a higher risk of placental anomalies compared with non-ART singleton pregnancies? A systematic review and meta-analysis. BJOG 126, 209–218 (2019).

4. Ceelen, M., van Weissenbruch, M.M., Vermeiden, J.P.W., van Leeuwen, F.E. & de Waal, H.A.D.V. Cardiometabolic differences in children born after in vitro fertilization: Follow-up study. J Clin Endocr Metab 93, 1682–1688 (2008).

5. Guo, X.Y. et al. Cardiovascular and metabolic profiles of offspring conceived by assisted reproductive technologies: a systematic review and meta-analysis. Fertility and Sterility 107, 622–631.e5. (2017).

6. Klemetti, R. et al. Puberty disorders among ART-conceived singletons: a Nordic register study from the CoNARTaS group. Hum Reprod 37, 2402–2411 (2022).

7. Lazaraviciute, G., Kauser, M., Bhattacharya, S., Haggarty, P. & Bhattacharya, S. A systematic review and meta-analysis of DNA methylation levels and imprinting disorders in children conceived by IVF/ICSI compared with children conceived spontaneously. Hum Reprod Update 20, 840–852 (2014).

8. Henningsen, A.A. et al. Imprinting disorders in children born after ART: a Nordic study from the CoNARTaS group. Hum Reprod 35, 1178–1184 (2020).

9. Rönö, K. et al. The neurodevelopmental morbidity of children born after assisted reproductive technology: a Nordic register study from the Committee of Nordic Assisted Reproductive Technology and Safety group. Fertility and Sterility 117, 1026–1037 (2022).

10. Maheshwari, A. et al. Is frozen embryo transfer better for mothers and babies? Can cumulative meta-analysis provide a definitive answer? Human Reproduction Update 24, 35–58 (2018).

11. Melamed, N., Choufani, S., Wilkins-Haug, L.E., Koren, G. & Weksberg, R. Comparison of genome-wide and gene-specific DNA methylation between ART and naturally conceived pregnancies. Epigenetics-Us 10, 474–483 (2015).

12. Estill, M.S. et al. Assisted reproductive technology alters deoxyribonucleic acid methylation profiles in bloodspots of newborn infants. Fertility and Sterility 106, 629–639.e10. (2016).

13. El Hajj, N., et al. DNA methylation signatures in cord blood of ICSI children. Hum Reprod 32, 1761–1769 (2017).

14. Novakovic, B. et al. Assisted reproductive technologies are associated with limited epigenetic variation at birth that largely resolves by adulthood. Nat Commun 10, 3922 (2019).

15. Tobi, E.W. et al. DNA methylation differences at birth after conception through ART. Hum Reprod 36, 248-259 (2021).

16. Yeung, E.H. et al. Conception by fertility treatment and offspring deoxyribonucleic acid methylation. Fertility and Sterility 116, 493–504 (2021).

17. Håberg, S.E. et al. DNA methylation in newborns conceived by assisted reproductive technology. Nat Commun 13, 1896 (2022).

18. Rahimi, S. et al. Capturing sex-specific and hypofertility-linked effects of assisted reproductive technologies on the cord blood DNA methylome. Clin Epigenetics 15, 82 (2023).

19. Litzky, J.F. et al. Placental imprinting variation associated with assisted reproductive technologies and subfertility. Epigenetics-Us 12, 653–661 (2017).

20. Xu, N. et al. Comparison of Genome-Wide and Gene-Specific DNA Methylation Profiling in First-Trimester Chorionic Villi From Pregnancies Conceived With Infertility Treatments. Reprod Sci 24, 996–1004 (2017).

21. Choufani, S. et al. Impact of assisted reproduction, infertility, sex and paternal factors on the placental DNA methylome. Hum Mol Genet 28, 372–385 (2019).

22. Mani, S. et al. Embryo cryopreservation leads to sex-specific DNA methylation perturbations in both human and mouse placentas. Hum Mol Genet 31, 3855–3872 (2022).

23. Mann, M.R.W. et al. Selective loss of imprinting in the placenta following preimplantation development in culture. Development 131, 3727–3735 (2004).

24. de Waal, E. et al. The cumulative effect of assisted reproduction procedures on placental development and epigenetic perturbations in a mouse model. Hum Mol Genet 24, 6975–6985 (2015).

25. Rahimi, S. et al. Moderate maternal folic acid supplementation ameliorates adverse embryonic and epigenetic outcomes associated with assisted reproduction in a mouse model. Hum Reprod 34, 851–862 (2019).

26. Ghosh, J., Coutifaris, C., Sapienza, C. & Mainigi, M. Global DNA methylation levels are altered by modifiable clinical manipulations in assisted reproductive technologies. Clin Epigenetics 9, 14 (2017).

27. Choux, C. et al. The epigenetic control of transposable elements and imprinted genes in newborns is affected by the mode of conception: ART versus spontaneous conception without underlying infertility. Hum Reprod 33, 331–340 (2018).

28. Barberet, J. et al. Do frozen embryo transfers modify the epigenetic control of imprinted genes and transposable elements in newborns compared with fresh embryo transfers and natural conceptions? Fertility and Sterility 116, 1468–1480 (2021).

29. Gong, S. et al. Genome-wide oxidative bisulfite sequencing identifies sex-specific methylation differences in the human placenta. Epigenetics-Us 13, 228–239 (2018).

30. Andrews, S.V., Yang, I.J., Froehlich, K., Oskotsky, T. & Sirota, M. Large-scale placenta DNA methylation integrated analysis reveals fetal sex-specific differentially methylated CpG sites and regions. Sci Rep-Uk 12, 9396 (2022).

31. Sandman, C.A., Glynn, L.M. & Davis, E.P. Is there a viability-vulnerability tradeoff? Sex differences in fetal programming. J Psychosom Res 75, 327–335 (2013).

32. Tarrade, A., Panchenko, P., Junien, C. & Gabory, A. Placental contribution to nutritional programming of health and diseases: epigenetics and sexual dimorphism. J Exp Biol 218, 50–58 (2015).

33. Kaartinen, N.M. et al. Male gender explains increased birthweight in children born after transfer of blastocysts. Hum Reprod 30, 2312–2320 (2015).

34. Litzky, J.F. et al. Effect of frozen/thawed embryo transfer on birthweight, macrosomia, and low birthweight rates in US singleton infants. Am J Obstet Gynecol 218, 433.e1–433.e10. (2018).

35. Coetzee, K., Ozgur, K., Bulut, H., Berkkanoglu, M. & Humaidan, P. Large-for-gestational age is male-gender dependent in artificial frozen embryo transfers cycles: a cohort study of 1295 singleton live births. Reprod Biomed Online 40, 134–141 (2020).

36. Sankilampi, U., Hannila, M.L., Saari, A., Gissler, M. & Dunkel, L. New population-based references for birth weight, length, and head circumference in singletons and twins from 23 to 43 gestation weeks. Ann Med 45, 446–454 (2013).

37. Gomes, L.G. et al. DLK1 Is a Novel Link Between Reproduction and Metabolism. J Clin Endocr Metab 104, 2112–2120 (2019).

38. Tan, J.H.L., Wollmann, H., van Pelt, A.M.M., Kaldis, P. & Messerschmidt, D.M. Infertility-Causing Haploinsufficiency Reveals TRIM28/KAP1 Requirement in Spermatogonia. Stem Cell Rep 14, 818–827 (2020).

39. Dalgaard, K. et al. Trim28 Haploinsufficiency Triggers Bi-stable Epigenetic Obesity. Cell 164, 353–364 (2016).

40. Li, R. et al. TRIM28 modulates nuclear receptor signaling to regulate uterine function. Nat Commun 14, 4605 (2023).

41. Bond, S.T. et al. Deletion of Trim28 in committed adipocytes promotes obesity but preserves glucose tolerance. Nat Commun 12, 74 (2021).

42. Domenga, V. et al. Notch3 is required for arterial identity and maturation of vascular smooth muscle cells. Gene Dev 18, 2730–2735 (2004).

43. Dietrich, B.V., K; Lackner, A. I.; Meinhardt, G; Koo, B-K.; Pollheimer, J.; Haider, S; Knöfler, M. NOTCH3 signalling controls human trophoblast stem cell expansion and differentiation. bioRxiv 2023.07.03.547490(2023).

44. Afshar, Y. et al. Notch1 mediates uterine stromal differentiation and is critical for complete decidualization in the mouse. Faseb J 26, 282–294 (2012).

45. Kim, J.Y. & Kim, Y.M. Acute Atherosis of the Uterine Spiral Arteries: Clinicopathologic Implications. J Pathol Transl Med 49, 462–471 (2015).

46. Dicke, A.K. et al. DDX3Y is likely the key spermatogenic factor in the AZFa region that contributes to human non-obstructive azoospermia. Commun Biol 6, 350 (2023).

47. Holmlund, H., Yamauchi, Y., Ruthig, V.A., Cocquet, J. & Ward, M.A. Return of the forgotten hero: the role of Y chromosome-encoded Zfy in male reproduction. Mol Hum Reprod 29, gaad025 (2023).

48. Anilkumar, T.R. et al. Expression of protocadherin 11Yb (PCDH11Yb) in seminal germ cells is correlated with fertility status in men. Reprod Fert Develop 29, 2100–2111 (2017).

49. Qin, Y.F. et al. Genetic variants in epoxide hydrolases modify the risk of oligozoospermia and asthenospermia in Han-Chinese population. Gene 510, 171–174 (2012).

50. Thelin, M.A. et al. Biological functions of iduronic acid in chondroitin/dermatan sulfate. Febs J 280, 2431–2446 (2013).

51. Eggermann, T. et al. Trans-acting genetic variants causing multilocus imprinting disturbance (MLID): common mechanisms and consequences. Clin Epigenetics 14, 41 (2022).

52. Li, X.J., et al. A Maternal-Zygotic Effect Gene, Zfp57, Maintains Both Maternal and Paternal Imprints. Dev Cell 15, 547–557 (2008).

54. Quenneville, S., et al. In Embryonic Stem Cells, ZFP57/KAP1 Recognize a Methylated Hexanucleotide to Affect Chromatin and DNA Methylation of Imprinting Control Regions. Mol Cell 44, 361-372 (2011).

54. Cortessis, V.K. et al. Comprehensive meta-analysis reveals association between multiple imprinting disorders and conception by assisted reproductive technology. J Assist Reprod Gen 35, 943–952 (2018).

56. Jirtle, R.L. Geneimprint. Imprinted Genes: by Species. https://www.geneimprint.com/site/genes-by-species. (2023).

56. Barberet, J. et al. DNA methylation profiles after ART during human lifespan: a systematic review and meta-analysis. Human Reproduction Update 28, 629–655 (2022).

57. Ochoa, E. et al. ImprintSeq, a novel tool to interrogate DNA methylation at human imprinted regions and diagnose multilocus imprinting disturbance. Genet Med 24, 463–474 (2022).

58. Hara, S., Terao, M., Tsuji-Hosokawa, A., Ogawa, Y. & Takada, S. Humanization of a tandem repeat in IG-DMR causes stochastic restoration of paternal imprinting at mouse Dlk1-Dio3 domain. Hum Mol Genet 30, 564–574 (2021).

59. Alexander, K.A., Wang, X., Shibata, M., Clark, A.G. & Garcia-Garcia, M.J. TRIM28 Controls Genomic Imprinting through Distinct Mechanisms during and after Early Genome-wide Reprogramming. Cell Rep 13, 1194–1205 (2015).

60. Lu, H.P. et al. TRIM28 Regulates Dlk1 Expression in Adipogenesis. Int J Mol Sci 21, 7245 (2020).

61. Davies, M.J. et al. Reproductive Technologies and the Risk of Birth Defects. New Engl J Med 366, 1803–1813 (2012).

62. Auvinen, P. et al. Chromatin modifier developmental pluripotency associated factor 4 (DPPA4) is a candidate gene for alcohol-induced developmental disorders. Bmc Med 20, 495 (2022).

63. Laufer, B.I. et al. Placenta and fetal brain share a neurodevelopmental disorder DNA methylation profile in a mouse model of prenatal PCB exposure. Cell Rep 38, 110442 (2022).

64. Zhu, Y.H. et al. Placental methylome reveals a 22q13.33 brain regulatory gene locus associated with autism. Genome Biol 23, 46 (2022).

65. Dumoulin, J.C. et al. Growth rate of human preimplantation embryos is sex dependent after ICSI but not after IVF. Hum Reprod 20, 484–91 (2005).

66. Kai, C.M. et al. Serum insulin-like growth factor-I (IGF-I) and growth in children born after assisted reproduction. J Clin Endocr Metab 91, 4352–4360 (2006).

67. Umapathy, A. et al. Mesenchymal Stem/Stromal Cells from the Placentae of Growth Restricted Pregnancies Are Poor Stimulators of Angiogenesis. Stem Cell Rev Rep 16, 557–568 (2020).

68. Boss, A.L., Chamley, L.W., Brooks, A.E.S. & James, J.L. Differences in human placental mesenchymal stromal cells may impair vascular function in FGR. Reproduction 162, 319–330 (2021).

69. Wang, Y.F. et al. TRIM28 regulates sprouting angiogenesis through VEGFR-DLL4-Notch signaling circuit. Faseb J 34, 14710–14724 (2020).

70. Mitterberger, M.C. et al. DLK1(PREF1) is a negative regulator of adipogenesis in CD105(+)/CD90(+)/CD34(+)/CD31(-)/FABP4(-) adipose-derived stromal cells from subcutaneous abdominal fat pats of adult women. Stem Cell Res 9, 35–48 (2012).

71. Vietor, I. et al. The negative adipogenesis regulator Dlk1 is transcriptionally regulated by Ifrd1 (TIS7) and translationally by its orthologue Ifrd2 (SKMc15). Elife 12, e88350 (2023).

72. Moon, Y.S. et al. Mice lacking paternally expressed Pref-1/Dlk1 display growth retardation and accelerated adiposity. Mol Cell Biol 22, 5585–5592 (2002).

73. Villena, J.A. et al. Resistance to High-Fat Diet-Induced Obesity but Exacerbated Insulin Resistance in Mice Overexpressing Preadipocyte Factor-1 (Pref-1) A New Model of Partial Lipodystrophy. Diabetes 57, 3258–3266 (2008).

74. Dauber, A. et al. Paternally Inherited DLK1 Deletion Associated With Familial Central Precocious Puberty. J Clin Endocr Metab 102, 1557–1567 (2017).

75. Ioannides, Y., Lokulo-Sodipe, K., Mackay, D.J.G., Davies, J.H. & Temple, I.K. Temple syndrome: improving the recognition of an underdiagnosed chromosome 14 imprinting disorder: an analysis of 51 published cases. J Med Genet 51, 495–501 (2014).

76. Mackay, D.J.G. et al. Hypomethylation of multiple imprinted loci in individuals with transient neonatal diabetes is associated with mutations in ZFP57. Nat Genet 40, 949–951 (2008).

77. Fauque, P. et al. Reproductive technologies, female infertility, and the risk of imprinting-related disorders. Clin Epigenetics 12, 191 (2020).

78. Mu, J. et al. Mutations in NLRP2 and NLRP5 cause female infertility characterised by early embryonic arrest. J Med Genet 56, 471–480 (2019).

79. Barker, D.J., Winter, P.D., Osmond, C., Margetts, B. & Simmonds, S.J. Weight in infancy and death from ischaemic heart disease. Lancet 2, 577–580 (1989).

80. Barker, D.J.P. The developmental origins of chronic adult disease. Acta Paediatr 93, 26–33 (2004).

81. Cleaton, M.A.M. et al. Fetus-derived DLK1 is required for maternal metabolic adaptations to pregnancy and is associated with fetal growth restriction. Nat Genet 48, 1473–1480 (2016).

82. Declercq, E. et al. Perinatal outcomes associated with assisted reproductive technology: the Massachusetts Outcomes Study of Assisted Reproductive Technologies (MOSART). Fertil Steril 103, 888–895 (2015).

83. Tian, Y. et al. ChAMP: updated methylation analysis pipeline for Illumina BeadChips. Bioinformatics 33, 3982–3984 (2017).

84. Wang, Y.C. et al. InterpolatedXY: a two-step strategy to normalize DNA methylation microarray data avoiding sex bias. Bioinformatics 38, 3950–3957 (2022).

85. Pidsley, R. et al. A data-driven approach to preprocessing Illumina 450K methylation array data. Bmc Genomics 14, 293 (2013).

86. Johnson, W.E., Li, C. & Rabinovic, A. Adjusting batch effects in microarray expression data using empirical Bayes methods. Biostatistics 8, 118–127 (2007).

87. Zhou, W.D., Laird, P.W. & Shen, H. Comprehensive characterization, annotation and innovative use of Infinium DNA methylation BeadChip probes. Nucleic Acids Res 45, e22 (2017).

88. Hansen, K. IlluminaHumanMethylationEPICanno.ilm10b4.hg19: Annotation for Illumina’s EPIC methylation arrays. R package version 0.6.0. (2017).

90. Illumina. https://www.illumina.com. (2023).

90. Ritchie, M.E. et al. limma powers differential expression analyses for RNA-sequencing and microarray studies. Nucleic Acids Res 43, e47 (2015).

91. Yuan, V. et al. Cell-specific characterization of the placental methylome. Bmc Genomics 22, 6 (2021).

92. Kolde, R., Martens, K., Lokk, K., Laur, S. & Vilo, J. seqlm: an MDL based method for identifying differentially methylated regions in high density methylation array data. Bioinformatics 32, 2604–2610 (2016).

93. Phipson, B., Maksimovic, J. & Oshlack, A. missMethyl: an R package for analyzing data from Illumina’s HumanMethylation450 platform. Bioinformatics 32, 286–288 (2016).

94. Li, S. et al. Genome-wide average DNA methylation is determined in utero. Int J Epidemiol 47, 908–916 (2018).

95. Zheng, Y.A. et al. Prediction of genome-wide DNA methylation in repetitive elements. Nucleic Acids Res 45, 8697–8711 (2017).

97. Zymo Research. Bisulfite Primer Seeker. https://zymoresearch.eu/pages/bisulfite-primer-seeker. (2023).

97. Bock, C. et al. BiQ analyzer: visualization and quality control for DNA methylation data from bisulfite sequencing. Bioinformatics 21, 4067–4068 (2005).

98. Macosko, E.Z. et al. Highly Parallel Genome-wide Expression Profiling of Individual Cells Using Nanoliter Droplets. Cell 161, 1202–1214 (2015).

99. Love, M.I., Huber, W. & Anders, S. Moderated estimation of fold change and dispersion for RNA-seq data with DESeq2. Genome Biol 15, 550 (2014).

100. Zhang, Y.Q., Parmigiani, G. & Johnson, W.E. ComBat-seq: batch effect adjustment for RNA-seq count data. Nar Genom Bioinform 2, lqaa078 (2020).

101. Wu, T.Z. et al. clusterProfiler 4.0: A universal enrichment tool for interpreting omics data. Innovation-Amsterdam 2, 100141 (2021).

102. Team, R.C. R: a language and environment for statistical computing. Vienna: R Foundation for Statistical Computing. (2023).

