## Supplementary Figures for "Genome-wide DNA methylation, imprinting, and gene expression in human placentas derived from Assisted Reproductive Technology"

**Figure S1:** Cell type composition in placental samples.

**Figure S2:** Comparison of cell type compositions between control and ART placental samples.

**Figure S3:** GWAM comparison between control and ART placentas.

**Figure S4:** Comparison of RE DNAm between control and ART placentas.

**Figure S5:** DNAm profiles of control, ART, and SF placentas at *DLK1-DIO3* ICR.

**Figure S6:** Parental infertility diagnoses of ART, IUI, and SF newborns.

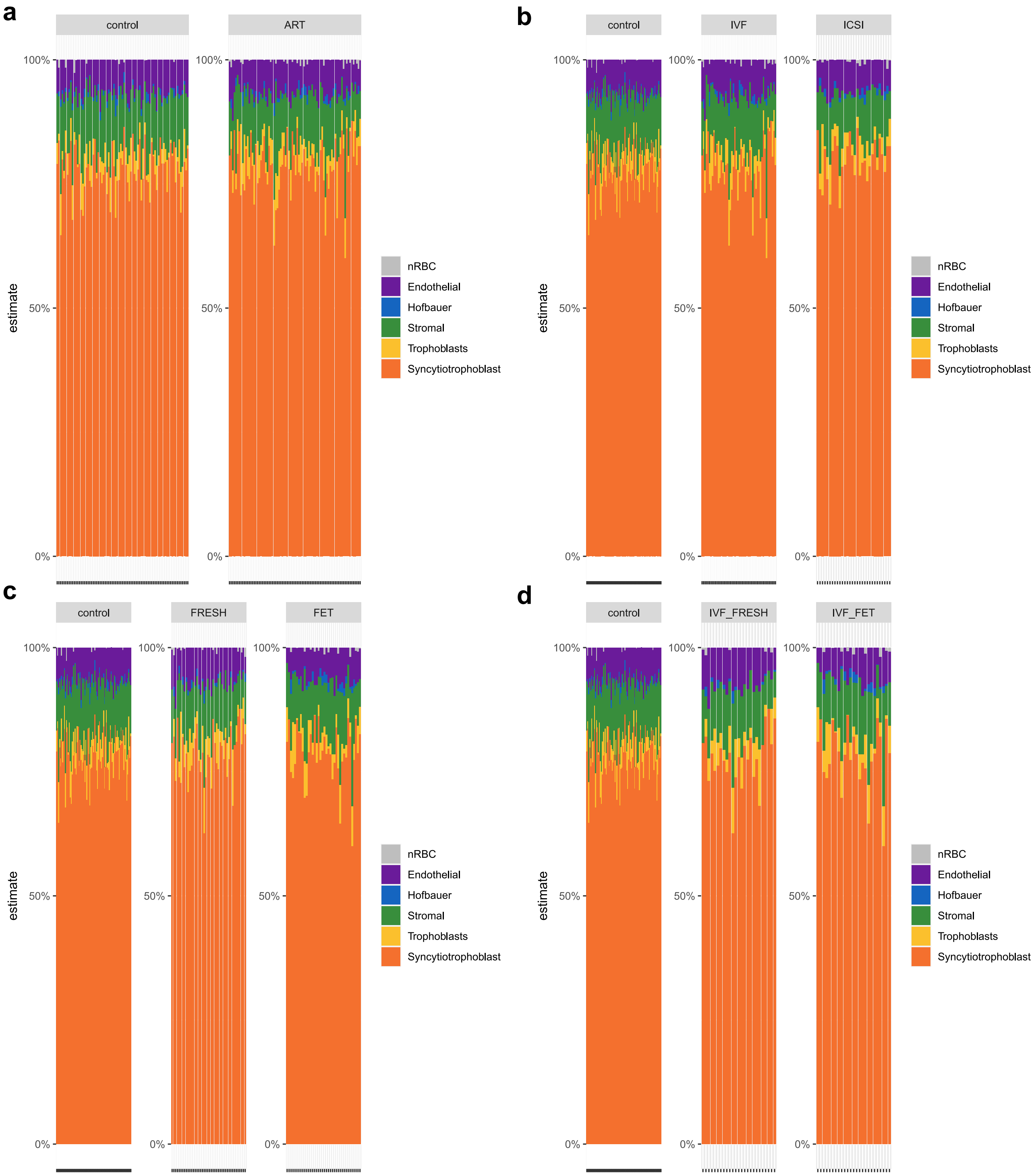


**Supplementary figure 1: Cell type composition in placental samples.** Cell type composition in **a** control and ART, **b** control, IVF, and ICSI, **c** control, FRESH, and FET as well as in **d** control, IVF-FRESH, and IVF-FET placental samples. Control *n* = 77, ART *n* = 80, IVF *n* = 50, ICSI *n* = 30, FRESH *n* = 42, FET *n* = 38, IVF-FRESH *n* = 25, and IVF-FET *n* = 25. nRBC: nucleated red blood cell.

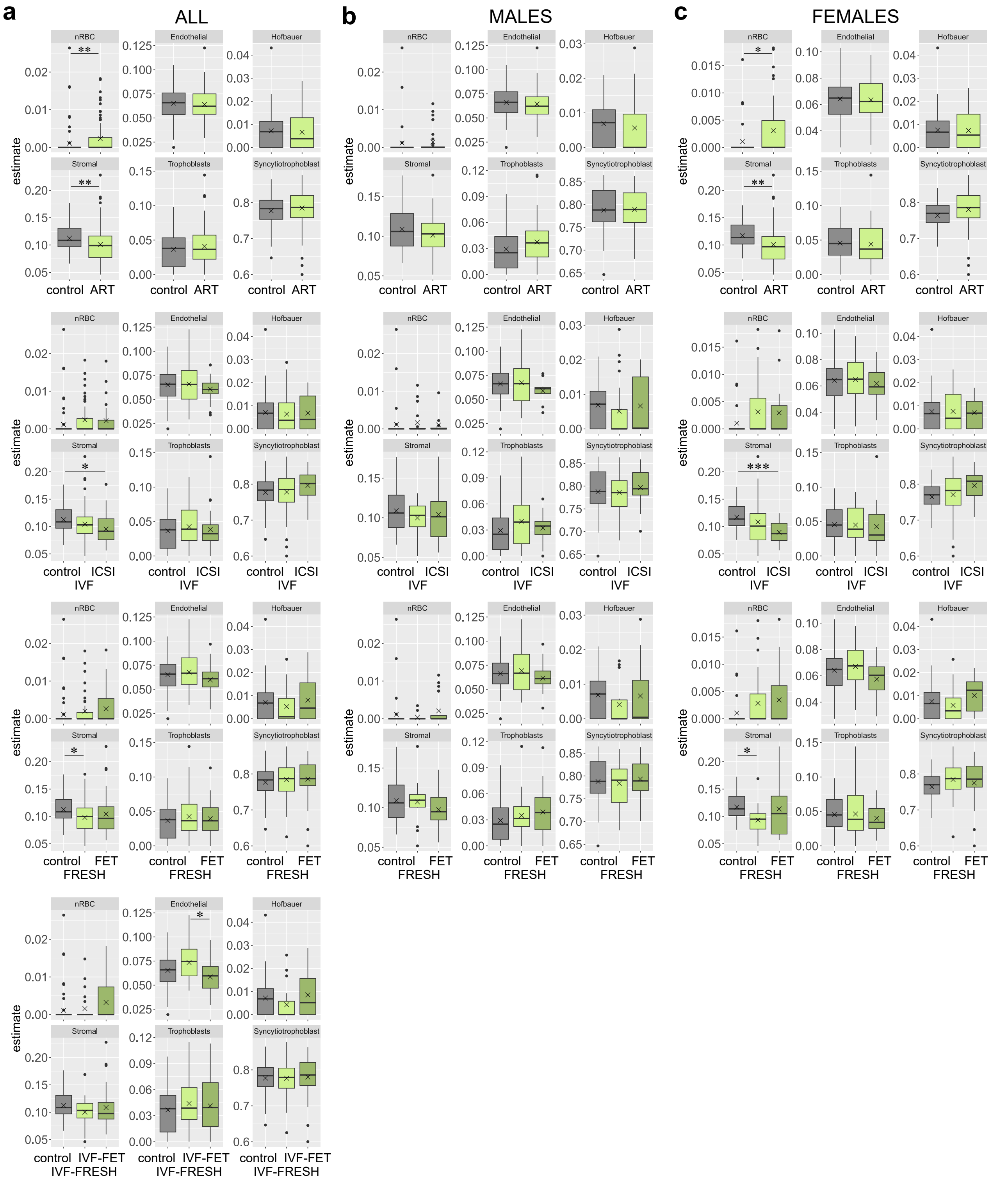


**Supplementary figure 2: Comparison of cell type compositions between control and ART placental samples.** Comparison of cell type compositions between **a** all, **b** male, and **c** female control, ART, and ART subgroup placental samples. **P* < 0.05 Wilcoxon Rank Sum Test or One-Way ANOVA followed by Tukey's HSD test, ***P* < 0.01, Wilcoxon Rank Sum Test, and ****P* < 0.001, Kruskal-Wallis test followed by Wilcoxon Rank Sum Exact test. Control *n* = 77 (42 males/35 females), ART *n* = 80 (36/44), IVF *n* = 50 (24/26), ICSI *n* = 30 (12/18), FRESH *n* = 42 (14/28), FET *n* = 38 (22/16), IVF-FRESH *n* = 25, and IVF-FET *n* = 25.

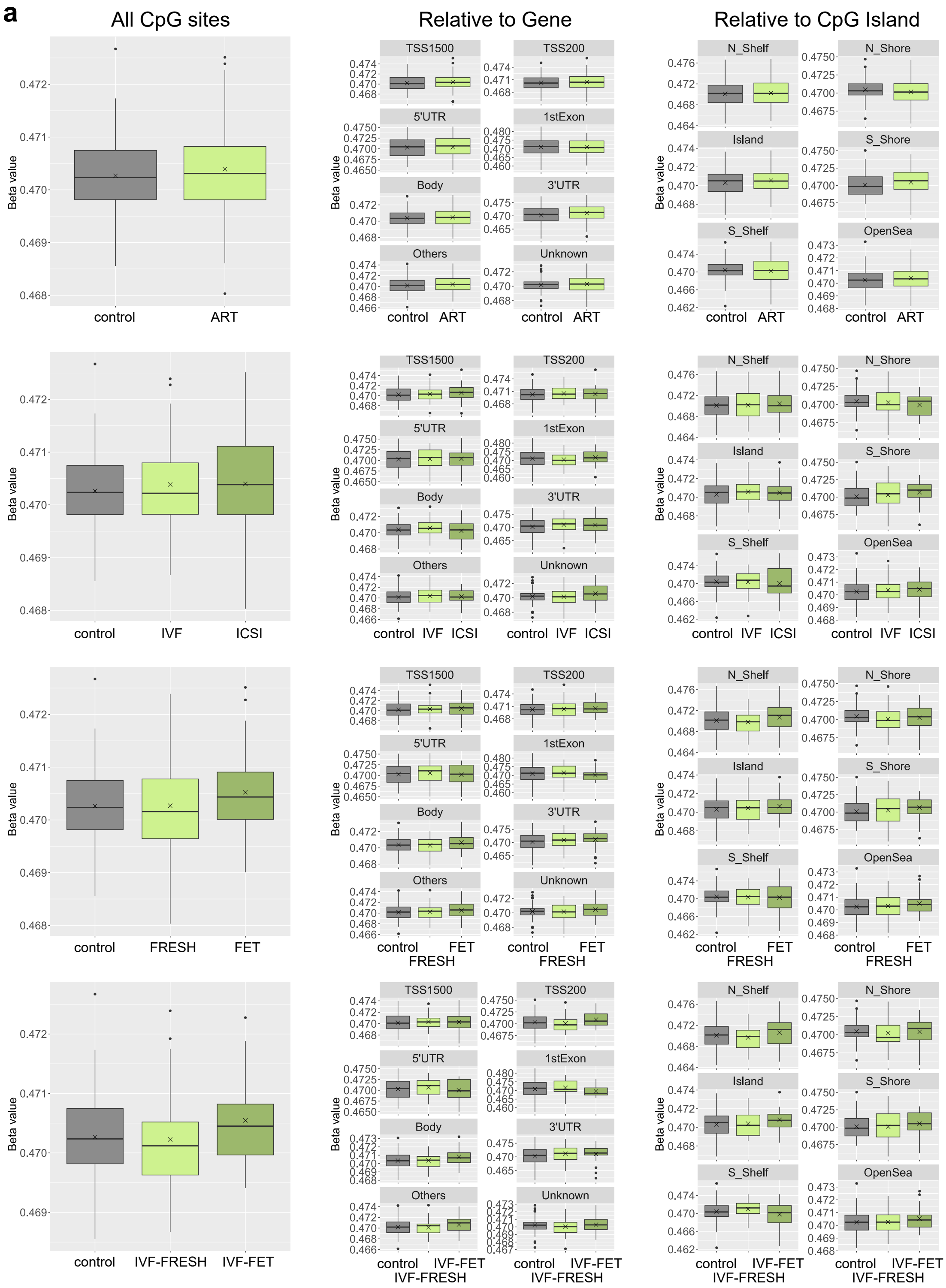
a
All CpG sites
Relative to Gene
Relative to CpG Island
Beta value
TSS1500
TSS200
5'UTR
1stExon
Body
3'UTR
Others
Unknown
N_Shelf
N_Shore
Island
S_Shore
S_Shelf
OpenSea
control
ART
IVF
ICSI
FRESH
FET
IVF-FRESH
IVF-FET

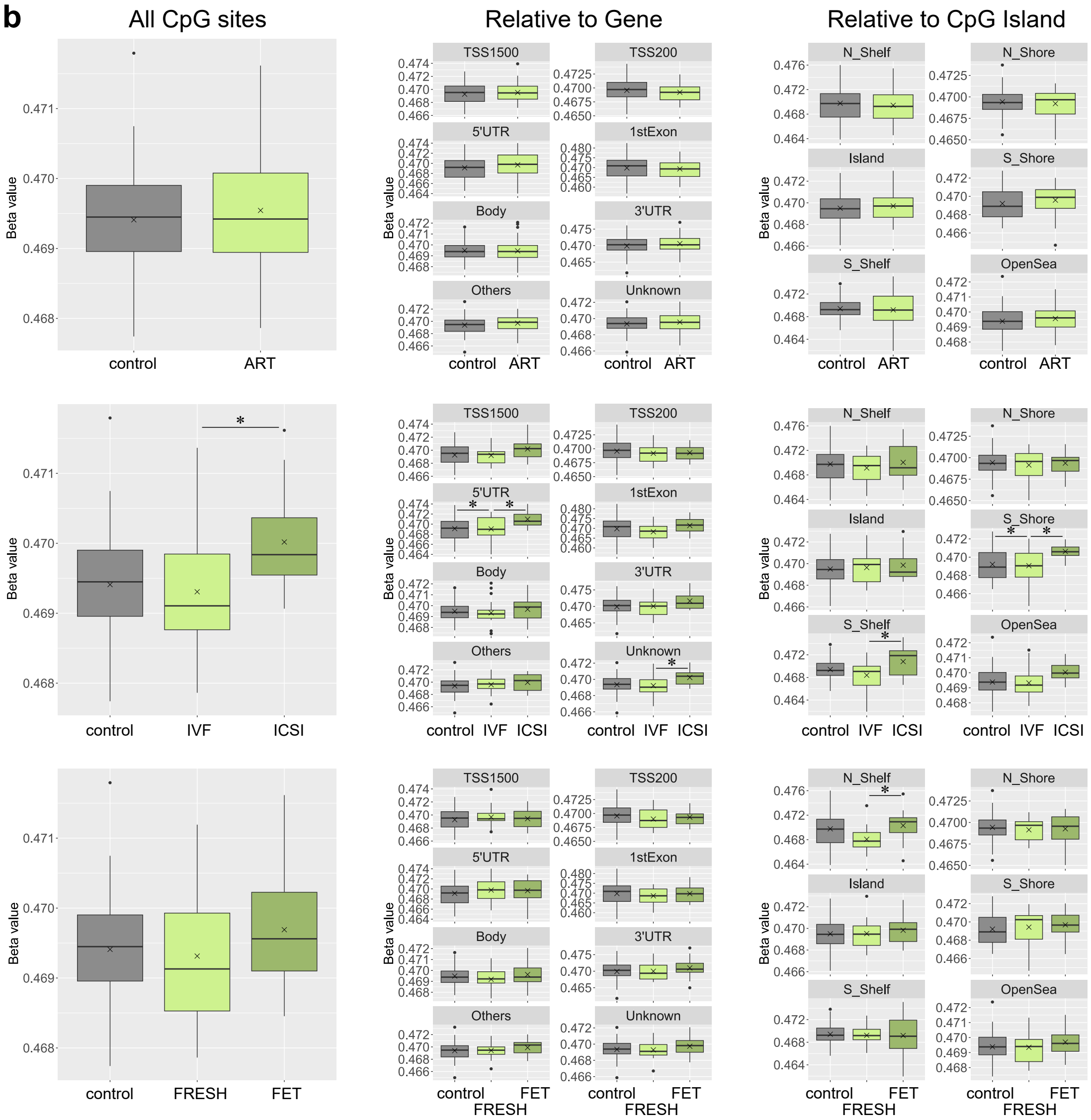
b
All CpG sites
Relative to Gene
Relative to CpG Island
Beta value
TSS1500
TSS200
5'UTR
1stExon
Body
3'UTR
Others
Unknown
N_Shelf
N_Shore
Island
S_Shore
S_Shelf
OpenSea
control
ART
IVF
ICSI
FRESH
FET

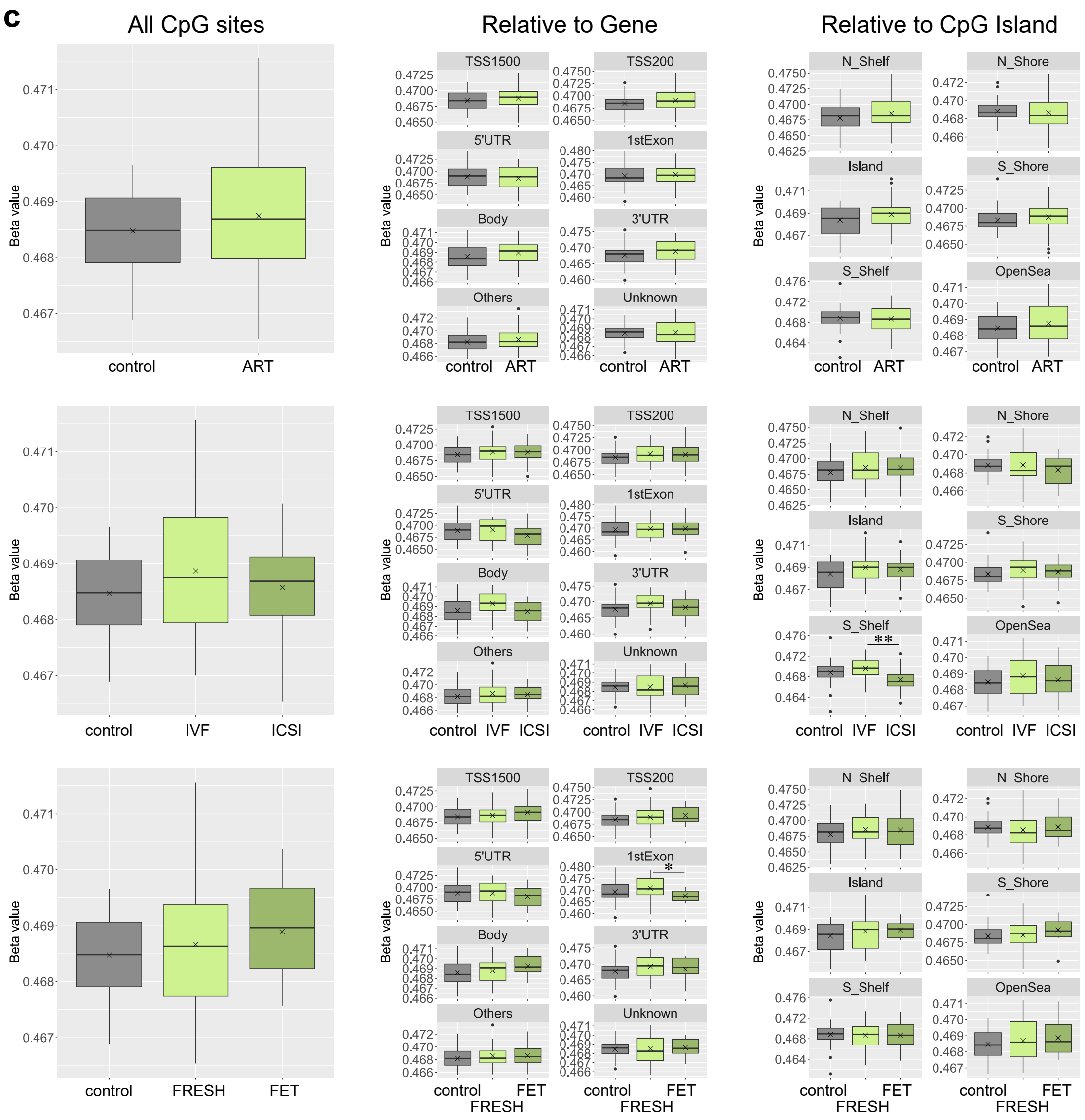


**Supplementary figure 3: GWAM comparison between control and ART placentas.** Comparison of DNAm in all probes, in relation to gene, and in relation to CpG island between **a** all, **b** male, and **c** female control, ART, and ART subgroup placentas. *$P$ < 0.05 and **$P$ < 0.01, One-Way ANOVA followed by Tukey's HSD test. Control $n$ = 77 (42 males/35 females), ART $n$ = 80 (36/44), IVF $n$ = 50 (24/26), ICSI $n$ = 30 (12/18), FRESH $n$ = 42 (14/28), FET $n$ = 38 (22/16), IVF-FRESH $n$ = 25, and IVF-FET $n$ = 25. TSS1500: 1500 bp upstream of transcription start site, TSS200: 200 bp upstream of TSS, UTR: untranslated region, N_shelf: north shelf, N_shore: north shore, S_shore: south shore, S_shelf: south shelf.

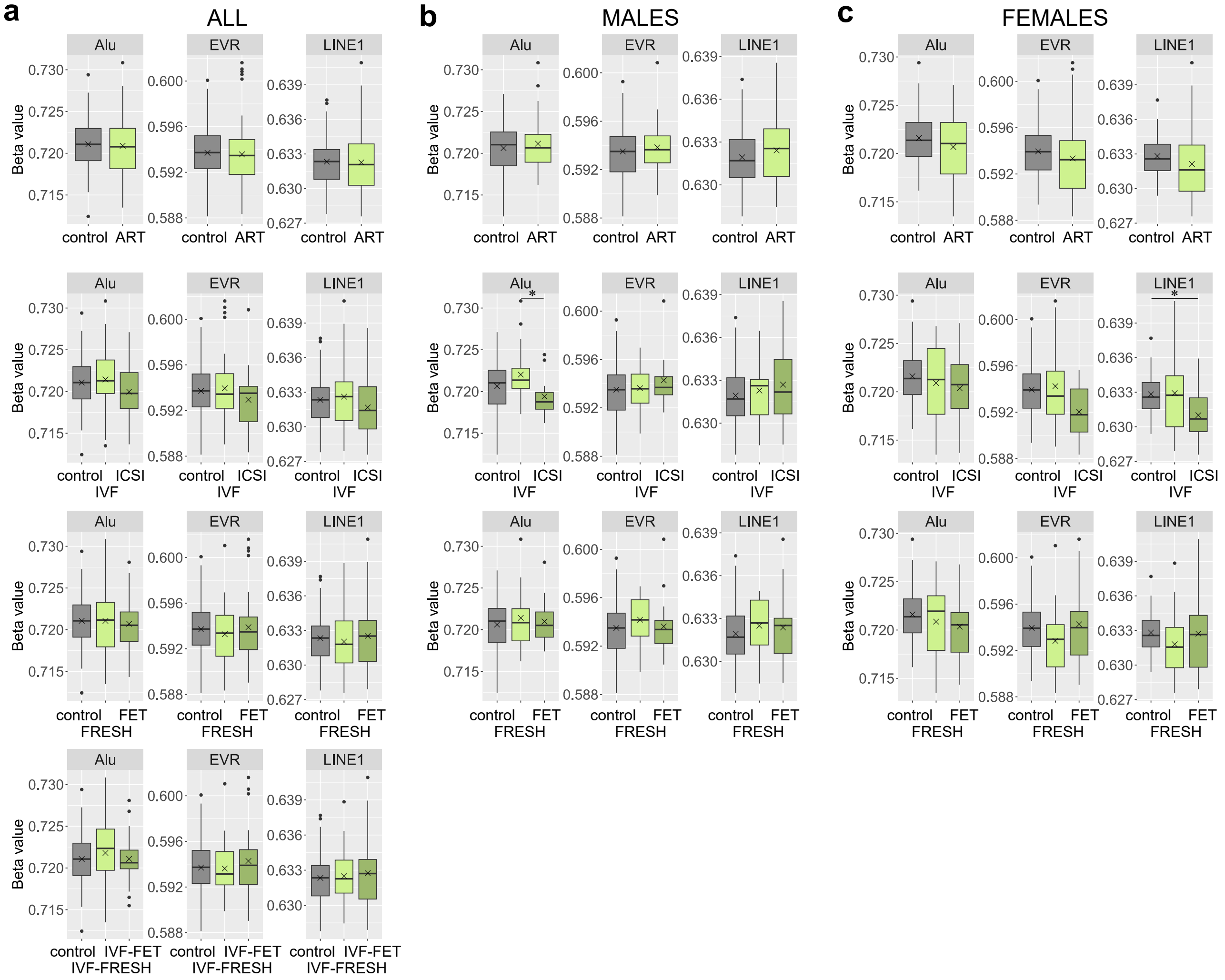


**Supplementary figure 4: Comparison of RE DNAm between control and ART placentas.** Comparison of DNAm in Alu, LINE1, and LTR repetitive regions between **a** all, **b** male, and **c** female control, ART, and ART subgroup placentas. **P* < 0.05, One-Way ANOVA followed by Tukey's HSD test. Control *n* = 77 (42 males/35 females), ART *n* = 80 (36/44), IVF *n* = 50 (24/26), ICSI *n* = 30 (12/18), FRESH *n* = 42 (14/28), FET *n* = 38 (22/16), IVF-FRESH *n* = 25, and IVF-FET *n* = 25.

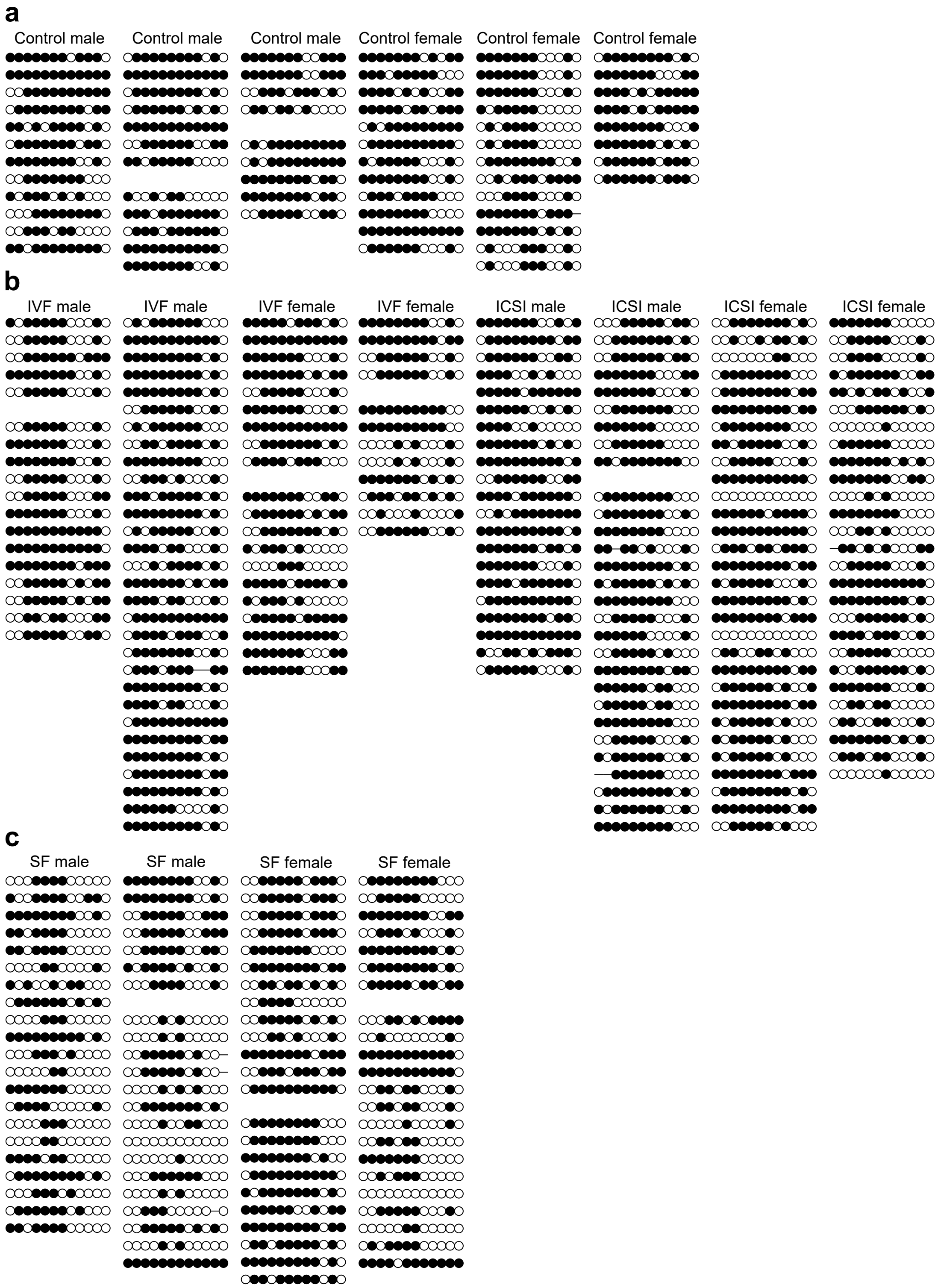


**Supplementary figure 5: DNAm profiles of control, ART, and SF placentas at *DLK1-DIO3* ICR.** Visualization of DNAm profiles at 12 CpG sites (chr14:101,277,375-101,277,826) in *DLK1-DIO3* ICR in **a** control ($n$ = 6), **b** IVF ($n$ = 4) and ICSI ($n$ = 4), as well as **c** in SF ($n$ = 4) male and female placentas. Paternal and maternal alleles are distinguished according to rs1884539(A/G) and rs75998174(A/G) polymorphisms when possible. Methylated and unmethylated CpGs are shown in black and white circles, respectively.

**a**

**b**

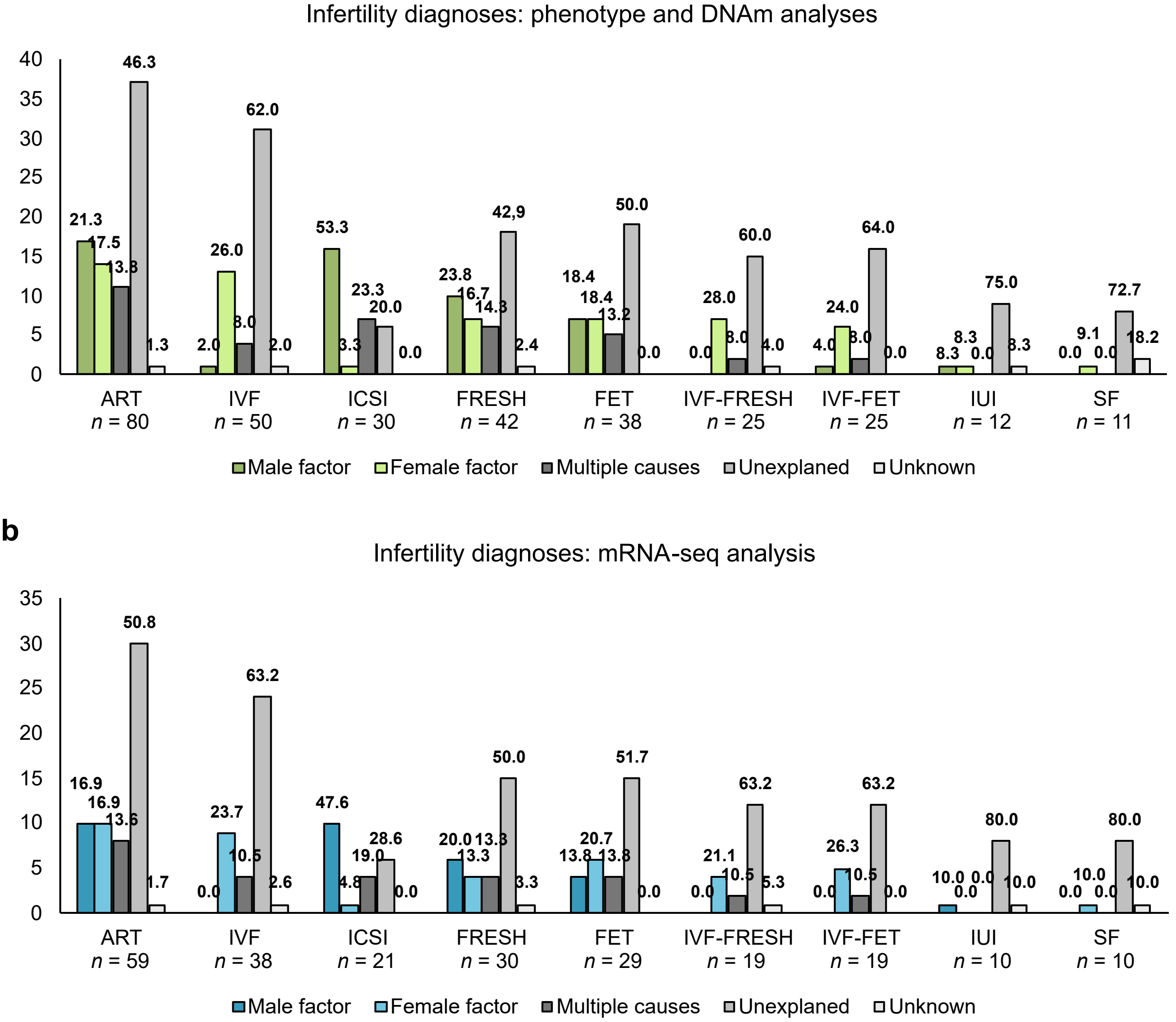


**Supplementary figure 6: Parental infertility diagnoses of ART, IUI, and SF newborns.** The proportions of categorized infertility diagnoses within study groups in **a** phenotype and genome-wide DNAm analysis, and in **b** genome-wide mRNA-seq analysis. The percentages of categorized diagnoses in each group are indicated above the bars. The proportions of the diagnoses in sex-specific analyses are not presented.
